# Defining Epidemiologically Relevant Units of Cholera Transmission in sub-Saharan Africa

**DOI:** 10.1101/2025.06.06.25329161

**Authors:** Bethany L. DiPrete, Javier Perez-Saez, Shirlee Wohl, Nathaniel Matteson, Seungwon Kim, Andrew S. Azman, Justin Lessler

## Abstract

Much remains unknown about cholera transmission dynamics in Africa. Applying both traditional phylogeographic techniques and novel methods for combining epidemiologic and genomic data, we identified 3-4 distinct ‘cholera transmission units’ within which cross-border cholera transmission has been more frequent. These units can be grouped into two major transmission clusters, one in Western Africa and bordering countries, and one spanning the Central, Eastern, and Southern regions. Simulation studies show that these clusters can be used to assess the risk of international spread of introduced cholera strains and identify heterogeneities in the speed of international spread based on outbreak location. Our methods further allow us to infer cholera incidence attributable to specific lineages in the absence of sequencing data. This work has clear implications for cholera control.

---

Cholera is an acute gastrointestinal infection caused by toxigenic *Vibrio cholerae* bacillus. Disease is characterized by watery diarrhea and rapid dehydration, which can rapidly progress to death if untreated. Despite a multitude of proven tools for cholera control, including improved water and sanitation infrastructure, vaccines, and effective treatment with oral rehydration salts, cholera continues to cause significant morbidity and mortality, with an estimated 1.3-4.0 million cases and 21,000-143,000 deaths annually (*1*). The disease is widespread in its ancestral home in countries surrounding the Bay of Bengal (*2–4*); however, much of the world’s cholera mortality is concentrated in sub-Saharan Africa (*5*).

Cholera has been endemic in sub-Saharan Africa since the 1960s when pandemic O1 El Tor *V. cholerae* was introduced from Southeast Asia (*2*). Incidence in the region tends to follow seasonal patterns (*6, 7*), but large, often off-season outbreaks are not uncommon and can have devastating impacts (*8, 9*). Natural disasters and social disruptions often trigger major cholera epidemics (*10, 11*), but do not explain cholera’s persistence and spread throughout the continent. To uncover the underlying drivers of cholera spread, scientists have turned to whole-genome sequencing of *V. cholerae* and found that there have been at least 17 introductions of O1 El Tor *V. cholerae* (lineages AFR1-AFR17) from Southeast Asia into Africa since 1970 (*12, 13*). Recent studies provide some insight into how cholera moves throughout the continent once introduced, finding evidence for frequent cross-border transmission between nearby countries, strain co-circulation within countries, and long-term persistence of lineages during epidemic lulls (*13–15*). However, these studies have been limited in their geographic and temporal scope, in part due to the limited and uneven coverage of available sequence data (**Fig. 1A**). Hence, questions remain about the geographic structure of cholera transmission within the continent, lineage persistence, and how newly introduced lineages are likely to spread.

**Figure 1.**
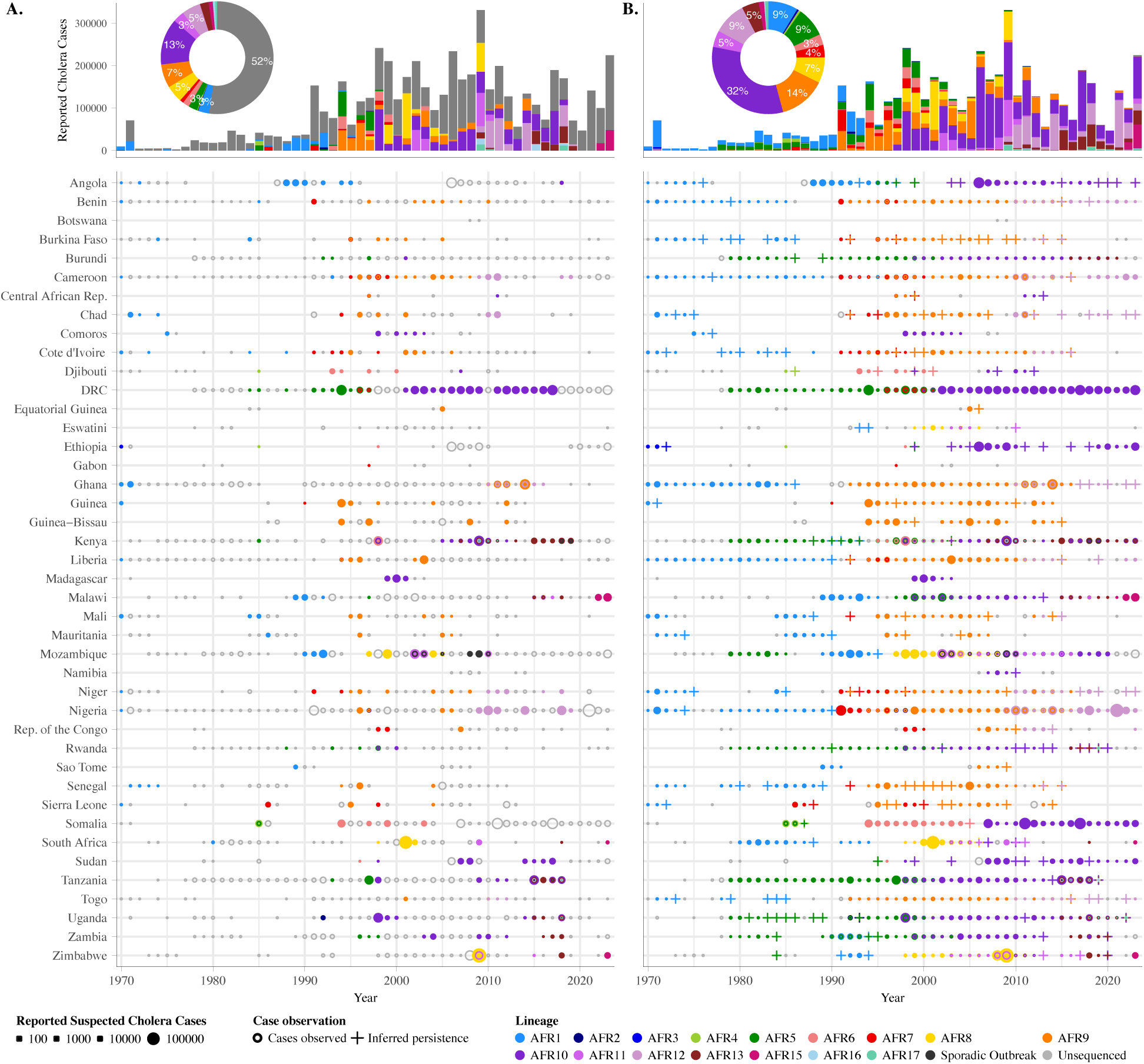
Reported cholera cases and lineages by country, 1970-2023. All sub-Saharan African countries are included. Sudan and South Sudan are geographically combined in analyses due to complexities in disentangling and modeling cases and infecting lineages before and after 2014. We recognize that the two countries are politically separate and may have different transmission dynamics. **(A)** Reported suspected cholera cases, depicted by the size of the circle, and detected lineages, indicated by the color of the circle. Multiple colors within a circle indicate >1 lineages detected through sequencing, whereas an empty grey circle indicates no sequences available. Bar graph displays total cases by year attributable to each lineage, and donut plot shows the proportion of cases attributable to each lineage overall. **(B)** Reported suspected cholera cases, depicted by the size of the circle. Detected lineages and lineages inferred through modeling are indicated by the color of the circle, where multiple colors within a circle indicate >1 lineages inferred or detected through sequencing. Crosses indicate inferred lineage presence when no cases were reported. Bar graph displays total cases by year attributable to a given observed or inferred lineage, and donut plot shows the proportion of cases attributable to each lineage overall.

## Phylogeographic Analysis

We used phylogeographic techniques to infer international cholera transition rates within Africa based on 1,772 third-wave (*16*) pandemic (AFR9 onwards) cholera sequences (**table S1**) collected from 36 African countries between 1989 and 2023 **(Fig. 2, 3A-B; see Methods)**. We observed the highest density and magnitude of geographic transitions between spatially proximal countries, with infrequent support for transitions between more distant locations (**Fig. 3A**), consistent with previous regional studies (*13–15, 17*). For instance, with every additional 1000 km distance between country centroids, the probability of a supported transition decreased by 88% (OR 0.12; 95% CI 0.08,0.18).

**Fig. 2.**
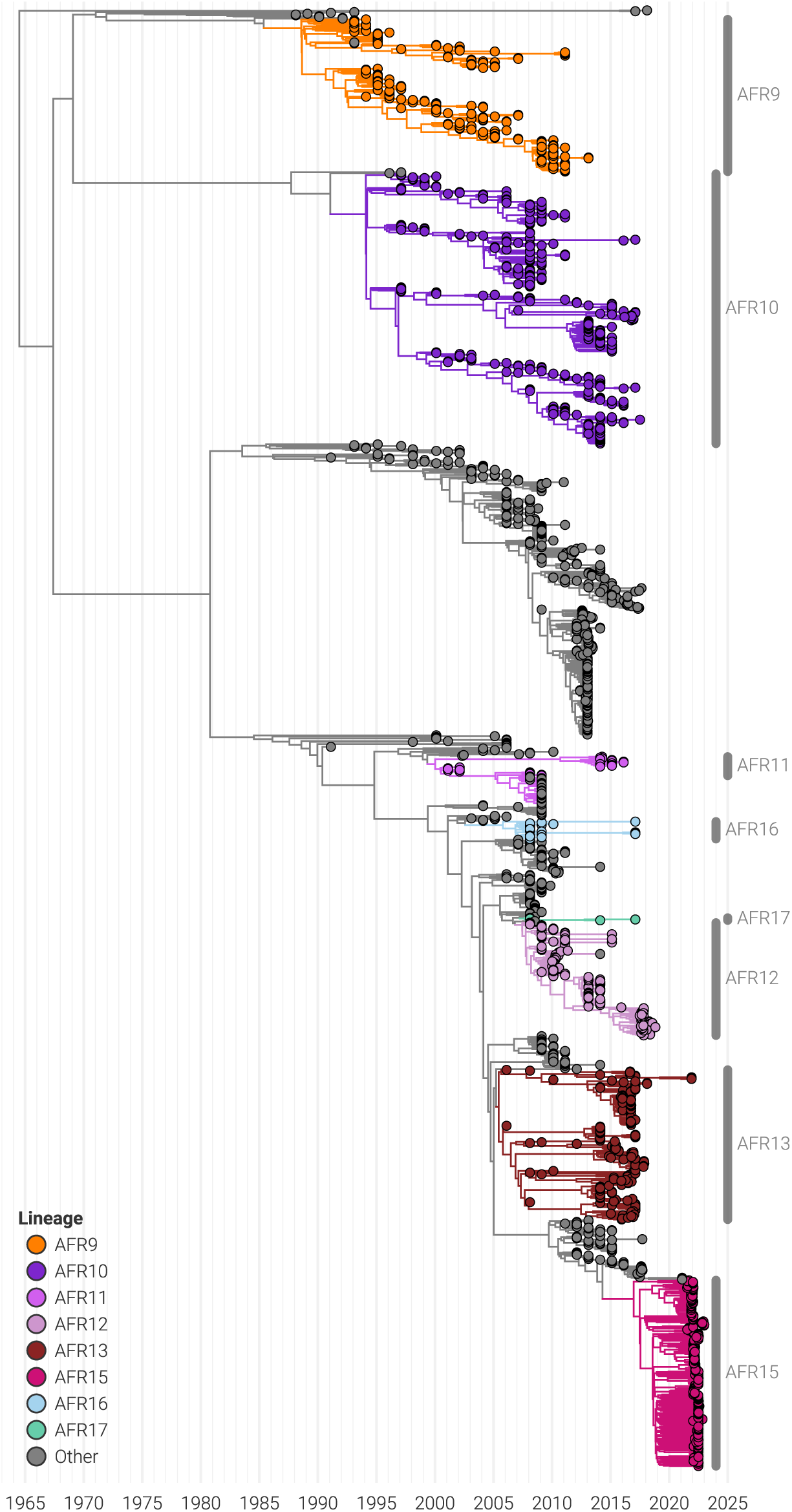
Phylogeny of third-wave seventh pandemic *V. cholerae* El Tor genomes. Maximum likelihood phylogeny of 1,772 third-wave genomes from 1989-2023 included in the phylogeographic analysis, with tip colored to indicate the inferred lineage.

**Figure 3.**
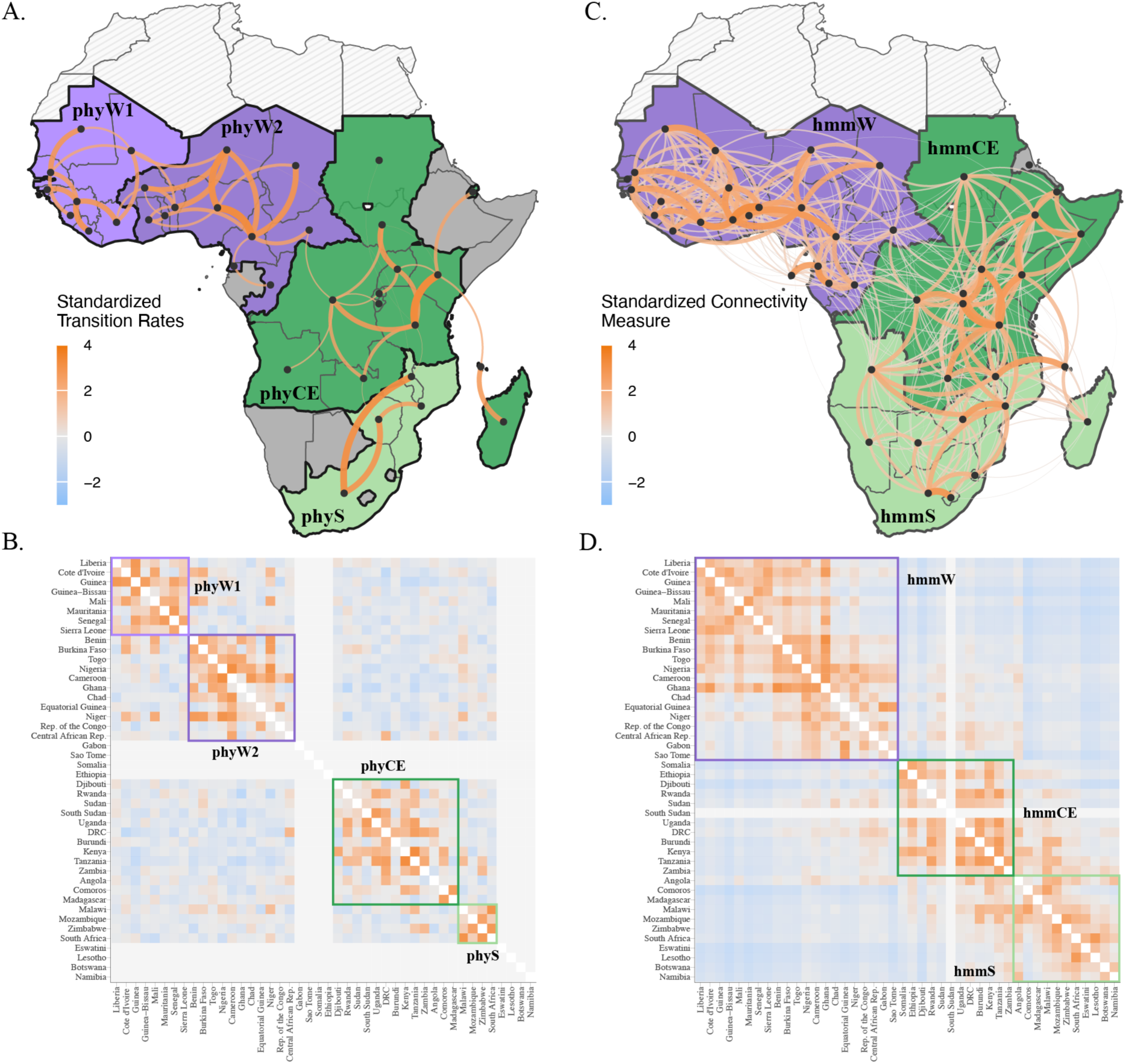
Inferred cholera transmission units and cross-country transmission in sub-Saharan Africa. **(A)** Results from phylogeographic analysis. Four identified cholera transmission units are shown by fill color (dark purple - phyW1, lighter purple - phyW2, dark green - phyCE, light green - phyS), with the color and thickness of edges representing the standardized transition rates between countries. Only edges with model support in 50% of runs are shown. Hashed grey indicates countries in Northern Africa that are not included in any analyses, whereas countries with the darker grey fill are not included in the phylogeographic analysis. **(B)** heat map of standardized transition rates between countries using the full network. **(C)** Results from HMM model. Three identified cholera transmission units are shown by fill color (purple - hmmW, dark green - hmmCE, light green - hmmS), with the color and and thickness of edges representing the standardized connectivity measure between countries. Only the top 50% of edges are plotted. Hashed grey indicates countries in Northern Africa that are not included in any analyses. **(D)** heat map of the standardized connectivity measure between countries using the full network.

We identified four distinct, but interconnected, ‘cholera transmission units’ defined by the probability of intra-versus extra-unit cholera transmission **(Fig. 3A-B)**. We first extracted non-overlapping communities from the network of transition rates between countries for each posterior sample using the Louvain method of community detection (*18*). Based on how consistently countries clustered together (**fig. S1A, Methods**), we then identified four cholera transmission units (*19*), two in Western Africa and neighboring countries (phyW1 and phyW2), one in Central and Eastern Africa (phyCE) and one in Southern Africa (phyS) (**Fig. 3A-B)**. Transition rates within cholera transmission units were, on average, 13.8 (95% HPD interval: 10.8, 17.4) times that between cholera transmission units. We were able to further group these transmission units into a Western cluster (including phyW1 and phyW2) and one spanning Central, Eastern, and Southern Africa (including phyCE and phyS) based on cross-unit transition rates (rates between phyW1 and phyW2 and between phyCE and phyS were 2.5 [1.6, 3.8] and 1.8 [0.7, 3.2] times the rates between the Western cluster and Central/Eastern/Southern cluster, respectively).

While this phylogeographic analysis provides important insights into cholera transmission patterns, it is limited by its reliance solely on sparse and unevenly distributed available sequence data (*20–22*). This analysis focused solely on third-wave data because, prior to 1989, sequence data were only available for 11% of country-year combinations where cholera was reported to the WHO. Even within the period covered, we have third-wave sequence data for only 25% of country-year combinations, and no cholera sequence data is available for 8 of 44 sub-Saharan African countries (**Fig. 1A**). Sparse coverage means that we are only able to infer infecting lineages based on local, contemporaneous, sequence data for 48% of African cholera cases since 1970. Hence, if we are to gain deeper insights into cholera transmission on the continent spanning the entirety of the seventh cholera pandemic, we must find a way to leverage the more widely available data on cholera incidence.

## Combining Epidemiologic and Lineage Data in a Hidden Markov Modelling Approach

We developed a novel modeling approach that leverages epidemiologic data and sequence-derived cholera lineages to infer missing cholera lineages and gain further insights into African transmission dynamics. We used a Hidden Markov Model (HMM) to synthesize incidence data from 1970-2023, covering 44 African countries with available lineage information from local, contemporaneous sequences (25% of country-years with reported cases). We modeled the underlying transmission and observation process of cholera to estimate the probability of lineage presence (its hidden presence state) across space and time, and the probability of lineage movement between pairs of countries (cross-border connectivity, **see Methods**).

### Inferring Presence, Prevalence, and Persistence of Infecting Lineages

Using this approach, we were able to infer infecting lineages for all reported cholera cases in sub-Saharan Africa **(Fig. 1B)**. We estimate that 45% of the cholera cases observed in Africa since 1970 (2.34 million cases) can be attributed to two lineages, AFR10 (32% [95% CI: 30, 35]) and AFR9 (14% [95% CI: 12, 15]), with no other lineage accounting for more than 10% of reported cases (**Fig. 1B)**. This analysis also gives further insight into local dynamics when sequence data is limited, e.g., suggesting the displacement of AFR5 by AFR10 in East Africa in the late 1990s and early 2000s was a trans-national phenomenon, and showing AFR10s persistence in these countries during periods with few cases and no sequence data (**Fig. 1B, S2**).

### Defining Cholera Transmission Units – HMM model

Using inferred connectivity from the HMM model, we identified three distinct cholera transmission units largely consistent with the phylogeographic analysis. Applying methods similar to those used to identify consensus clusters above to inferred connectivity measures (*18, 19*), we identified three cholera transmission units (**Fig. 3C-D, S1B**): one encompassing Western Africa and bordering countries (hmmW), a second spanning Central and Eastern Africa (hmmCE), and a third largely in Southern Africa (hmmS). As with the phylogeographic analysis, these cholera transmission units further grouped into two clusters, a Western cluster (hmmW) and one spanning Central, Eastern, and Southern Africa (hmmCE and hmmS). These major clusters were identical to those found in the phylogeographic analysis, while the individual cholera transmission units only differed in there being one (hmmW), versus two (phyW1, phyW2) Western transmission units and the assignment of Angola, Madagascar, and Comoros to the Southern (hmmS) rather than the Central/Eastern (hmmCE) transmission unit. Lineage movement between countries was 8.5 times (95% CI 6.9, 10.7) more likely within cholera transmission units (vs. between). The consistency of these results across methods affirms the distinctness of the Western cholera epidemic from the rest of the continent and shows the consistency of transmission patterns over time (clusters were similar when data were subset to 1990 onwards and 2000 onwards; see **fig. S3**).

### Simulating Onward Spread of Cholera

Simulations of the spread of newly introduced lineages made possible by the HMM model further support the importance of cholera transmission units. We used the HMM model to simulate onward spread from 2,000 introductions of cholera into each of 44 countries (sampling parameters from the posterior for each, **see Methods**). We found that within the first year after a simulated introduction into a single country, countries within the same cholera transmission unit were 10.7 times (95% CI 10.5, 10.9) more likely to experience a subsequent cholera outbreak compared to countries outside of that cholera transmission unit (**Fig. 4, S4**). Similarly, the average time to arrival of a newly introduced strain to countries within the same transmission unit was 3 years lower (95% CI −3.0, −2.9) than the average time to arrival for countries outside of that unit (**Fig. 5, S5**). Transmission between the Central/Eastern cholera transmission unit (hmmCE) and the Southern transmission unit (hmmS) was also more likely than between those units and the Western transmission unit (hmmW). For instance, an introduction in hmmCE is 4.5 times (95% CI 4.3, 4.8) more likely to cause an outbreak in hmmS in the next year versus causing one in hmmW.

**Fig. 4.**
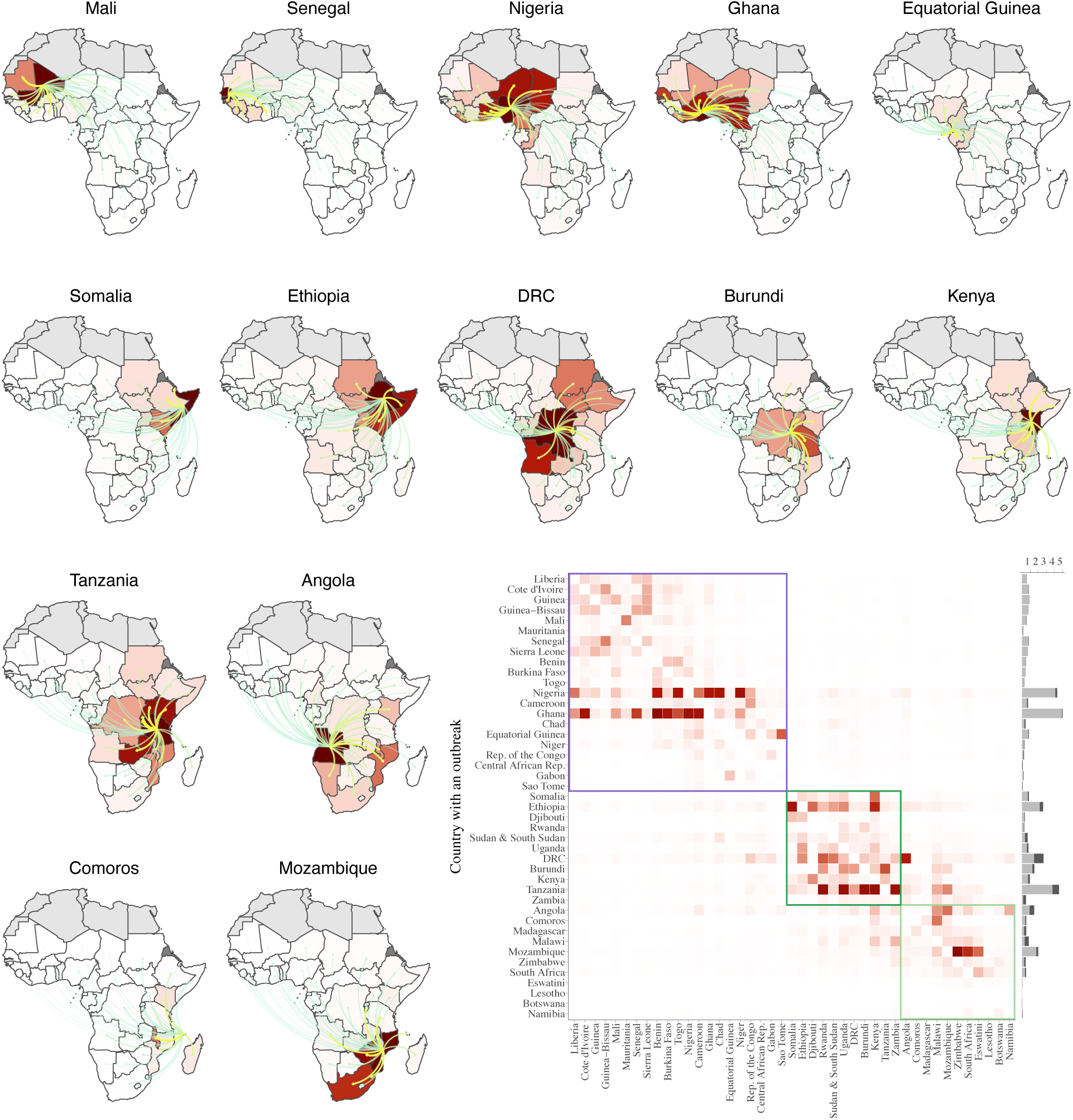
One-year risk of cholera outbreak by country of introduction. Simulated one-year risk of cholera outbreaks from 2,000 introductions of cholera into each of 44 countries (sampling parameters from the posterior for each), with darker red indicating higher risk of an outbreak in the following year. Edges indicate the asymmetrical strength of connectivity between the country of introduction and the rest of the continent. Maps are shown for a subset of countries with highest potential for seeding outbreaks. The heatmap displays outbreak risk across all countries, with the side panel showing the expected number of additional outbreaks caused within (light grey) and between (dark grey) transmission units by country of introduction.

**Fig. 5.**
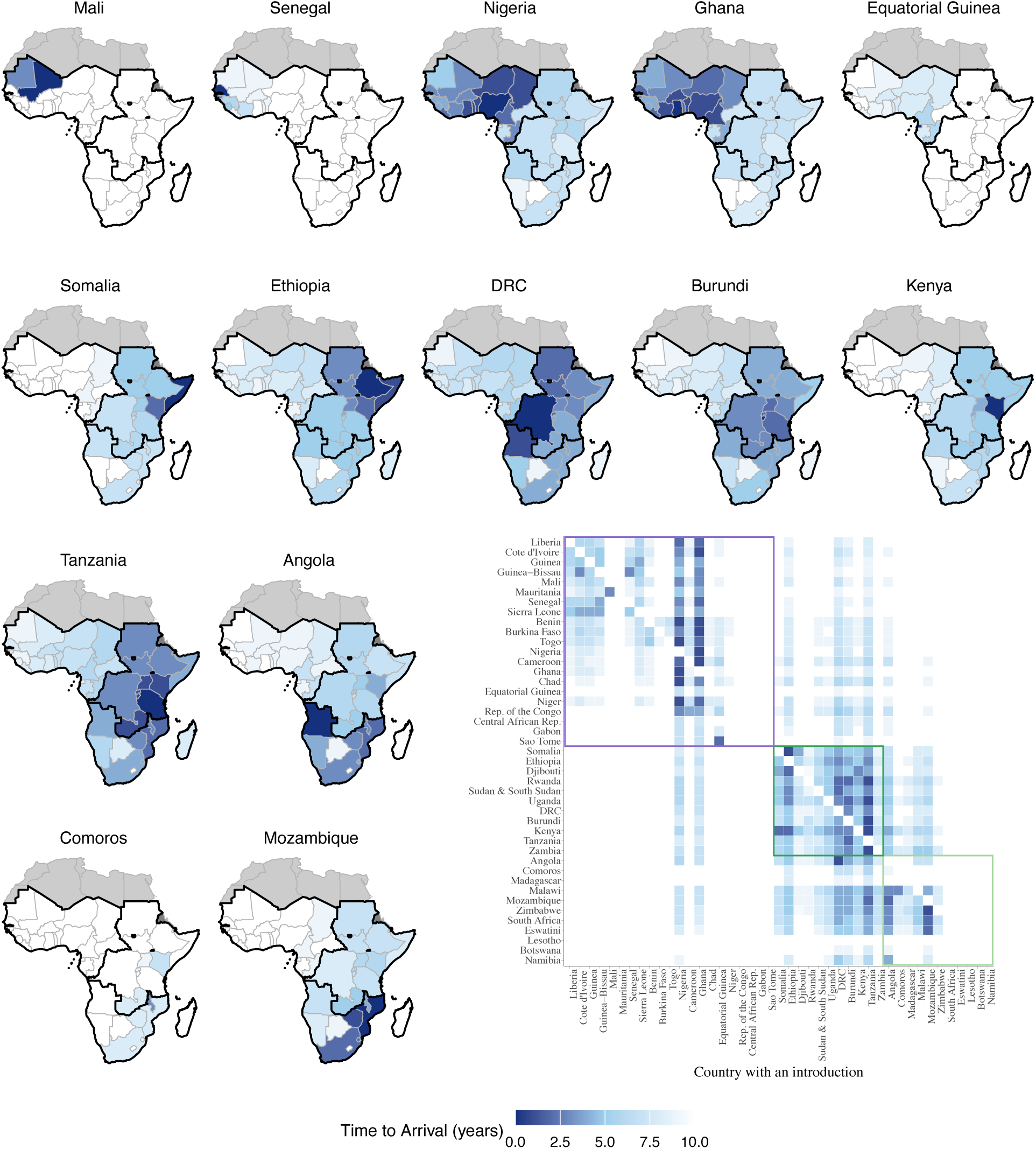
Speed of cholera spread by country of introduction. Simulated downstream time to arrival of cholera from 2,000 introductions of cholera into each of 44 countries (sampling parameters from the posterior for each), with darker blue indicating higher risk of an outbreak in the following year. Maps are shown for a subset of countries with highest potential for seeding outbreaks, whereas the heatmap shows time to arrival across all countries by country of introduction.

Even within transmission units, there are stark differences in the predicted extent and speed of spread from each country **(Fig. 4, 5, table S2)**. For example, an introduction into Ghana would be expected to cause outbreaks in 4.9 other countries within the Western cholera transmission unit (hmmW) within one year and be expected to reach at least 50% of the countries within that unit within 5.5 years. Similarly, an introduction into Nigeria would be expected to cause outbreaks in 4.1 countries in hmmW. However, an introduction into Senegal would only be expected to cause outbreaks in 0.8 countries in hmmW within a year and would take >10 years to spread across even a quarter of the cholera transmission unit. Likewise, some countries are more prone to spreading cholera between cholera transmission units than others. For instance, an introduction into Tanzania (in hmmCE) would be expected to cause 0.8 additional outbreaks in a different cholera transmission unit within a year (0.1 in hmmW, 0.7 in hmmS), while neighboring Kenya would only be expected to cause 0.2 such outbreaks (0.1 in hmmW, 0.2 in hmmS).

These results enrich our understanding of cholera transmission within Africa and have clear implications for assessing cholera risk and thinking about control. The existence of cholera transmission units provides a framework for determining what countries are most at risk from cholera outbreaks elsewhere in the continent and the natural grouping of countries over which we can coordinate cholera control. For instance, Cameroon might benefit more from coordinating cholera control efforts with Ghana (also in hmmW) than they would with the Democratic Republic of the Congo (in hmmCE), despite being geographically closer to the latter. Likewise, in response to a large outbreak of cholera in Angola (hmmS), prophylactic vaccination would be better targeted in Mozambique (hmmS) than the Republic of the Congo (hmmW). Our speed of spread simulations, though focused on newly introduced strains, provide some insight into country-to-country variation in the international spread of cholera resulting from outbreaks of all types. They suggest that outbreaks in some countries (e.g., Mozambique, DRC) are more likely to seed outbreaks elsewhere on the continent, while those in others (e.g., Senegal, Comoros) are likely to remain self-contained. This may help to further refine risk assessment for the distribution of prophylactic vaccines from the global stockpile and other interventions.

Our analysis of cholera transmission dynamics is not without limitations. As noted, the phylogeographic analysis was limited by data sparsity. Though the HMM modeling approach allowed us to infer much of this missing data, its inferences were also limited by data availability (as evidenced by improved performance in validation experiments in later years, see below), and came at the expense of losing valuable information on the genetic relationships between outbreaks of the same lineage. Further, inferences were based on reported suspected cholera cases, which are known to be an imperfect measure of true incidence (*1*). The use of country-level data likely obscures critical aspects of cholera dynamics. This may be particularly true in the DRC, where cholera epidemics in the east and west of the country are known to be relatively independent (*23*). Further, due to complexities in disentangling and modeling cases and lineages in Sudan and South Sudan before and after 2014, we geographically combined them for HMM analyses, while recognizing that the two countries are fully politically separate and may have different transmission dynamics. Our analysis takes no explicit account of immunity or strain-strain competition, which may play important roles in the spread of lineages and lineage replacement. Finally, our HMM analysis covered an extended time period over which cholera dynamics may have changed on the continent. This appears to only have a marginal effect as the results are largely consistent when we subset data to 1990 or 2000 onwards (**fig. S3, S6**), but may explain some of the differences between the phylogeographic and HMM analyses.

Despite these limitations, validation experiments show the robustness of our modeling approach. In cross-validation experiments where we removed 20% of detected lineages across countries and years (**see Methods**), the HMM model could infer detected lineages with 71% (SD = 6) recall (**table S3**). Later, more densely sampled lineages were recovered more consistently (83-87% recall for lineages AFR9-AFR13). If we presume clonal outbreaks (unless co-circulation is observed), precision was 60% (SD = 2); however, undetected co-circulation cannot be ruled out (and is assumed to be possible throughout the analysis), hence this may overestimate error (precision assuming nonclonal outbreaks: 82%, SD = 2). In simulation experiments where we compared the simulated time until a lineage would reach a country based on the country of introduction with observed arrival times (**fig. S7, see Methods**), we found moderate agreement between predicted and observed arrival times, both when the latter was based on the first observation of a lineage (Pearson *r*=0.44) and when based on the inferred arrival times from the full model fit (*r*=0.52). Again, predictions were more accurate for later lineages (*r*=0.82 for observed lineages). An advantage of this analysis is that it is based on actual cholera transmission patterns and not inferred from external sources.

This work illustrates the power of combining molecular and epidemiologic data to gain consequential insights into disease ecology (*17*). Phylogeographic analyses based on full genome sequences can provide a detailed understanding of the complex relationships between pathogen lineages, thereby revealing the ecologic and epidemiologic processes that create them. However, the sequences underlying these analyses will almost always be a biased and incomplete sample of the underlying pathogen population; hence may not give us a complete or fully accurate picture. By pairing phylogeographic analysis with complementary inferential methods that leverage both (less detailed) molecular relationships and epidemiologic data, we were able to gain deeper insights into cholera transmission in Africa. Our observations of clearly defined cholera transmission units, as well as inferences about lineage persistence and onward spread, can both inform public health action and drive further scientific enquiry. We also hope that this work can serve as a template for tighter integration of molecular and epidemiologic data in the study of other pathogens.

## Data Availability

Accession numbers and metadata for publicly available sequences used in this analysis are listed in Supplementary Materials (table S1). WHO annual case data by country is available upon request. . Code used in HMM model fitting and postestimation will be made available on GitHub prior to publication.

## Author contributions

Conceptualization: ASA, JL

Methodology: JL, JPS, BLD, ASA, SW, NM

Analysis: BLD, NM

Visualization: BLD, NM

Funding acquisition: ASA, JL

Supervision: ASA, JL

Writing – original draft: BLD, JL

Writing – review & editing: BLD, JL, JPS, SW, NM, ASA, SK

## Data and materials availability

Accession numbers and metadata for publicly available sequences used in this analysis are listed in Supplementary Materials (table. S1). WHO annual case data by country is available upon request. Code used in HMM model fitting and postestimation will be made available on GitHub prior to publication.

## Supplementary Materials

Materials and Methods

Figs. S1 to S7

Tables S1 to S3

References

## References and Notes

1. M. Ali, A. R. Nelson, A. L. Lopez, D. A. Sack, Updated global burden of cholera in endemic countries. PLoS Negl. Trop. Dis. 9, e0003832 (2015).

2. D. A. Sack, A. K. Debes, J. Ateudjieu, G. Bwire, M. Ali, M. C. Ngwa, J. Mwaba, R. Chilengi, C. C. Orach, W. Boru, A. A. Mohamed, M. Ram, C. M. George, O. C. Stine, Contrasting epidemiology of cholera in Bangladesh and Africa. J. Infect. Dis. 224, S701–S709 (2021).

3. J. Deen, M. A. Mengel, J. D. Clemens, Epidemiology of cholera. Vaccine 38 Suppl 1, A31–A40 (2020).

4. A. Barton, M. H. Afrad, A. Taylor-Brown, N. Singh, C. Thakur, T. Islam, S. I. A. Rahman, M. Akhtar, Y. A. Begum, T. R. Bhuiyan, A. I. Khan, N. Taneja, N. R. Thomson, F. Qadri, Evolution of Pandemic Cholera at its Global Source, medRxiv (2025) p. 2025.02.03.25321585.

5. M. A. Mengel, I. Delrieu, L. Heyerdahl, B. D. Gessner, Cholera outbreaks in Africa. Curr. Top. Microbiol. Immunol. 379, 117–144 (2014).

6. J. Perez-Saez, J. Lessler, E. C. Lee, F. J. Luquero, E. B. Malembaka, F. Finger, J. P. Langa, S. Yennan, B. Zaitchik, A. S. Azman, The seasonality of cholera in sub-Saharan Africa: a statistical modelling study. Lancet Glob Health 10, e831–e839 (2022).

7. S. M. Moore, A. S. Azman, B. F. Zaitchik, E. D. Mintz, J. Brunkard, D. Legros, A. Hill, H. McKay, F. J. Luquero, D. Olson, J. Lessler, El Niño and the shifting geography of cholera in Africa. Proc. Natl. Acad. Sci. U. S. A. 114, 4436–4441 (2017).

8. World Health Organization, “Disease Outbreak News; Cholera - Malawi” (2023); https://www-who-int.libproxy.lib.unc.edu/emergencies/disease-outbreak-news/item/2022-DON435.

9. J. Wise, Cholera: Malawi is in grip of its deadliest outbreak. BMJ 380, 328 (2023).

10. G. E. C. Charnley, K. Jean, I. Kelman, K. A. M. Gaythorpe, K. A. Murray, Association between conflict and cholera in Nigeria and the Democratic Republic of the Congo. Emerg. Infect. Dis. 28, 2472–2481 (2022).

11. Y. Ali, E. E. Siddig, A. Ahmed, Resurgence of cholera amidst ongoing war in Sudan. Lancet 404, 1724–1725 (2024).

12. F.-X. Weill, D. Domman, E. Njamkepo, C. Tarr, J. Rauzier, N. Fawal, K. H. Keddy, H. Salje, S. Moore, A. K. Mukhopadhyay, R. Bercion, F. J. Luquero, A. Ngandjio, M. Dosso, E. Monakhova, B. Garin, C. Bouchier, C. Pazzani, A. Mutreja, R. Grunow, F. Sidikou, L. Bonte, S. Breurec, M. Damian, B.-M. Njanpop-Lafourcade, G. Sapriel, A.-L. Page, M. Hamze, M. Henkens, G. Chowdhury, M. Mengel, J.-L. Koeck, J.-M. Fournier, G. Dougan, P. A. D. Grimont, J. Parkhill, K. E. Holt, R. Piarroux, T. Ramamurthy, M.-L. Quilici, N. R. Thomson, Genomic history of the seventh pandemic of cholera in Africa. Science 358, 785–789 (2017).

13. S. Xiao, A. Abade, W. Boru, W. Kasambara, J. Mwaba, F. Ongole, M. Mmanywa, N. S. Trovão, R. Chilengi, G. Kwenda, C. G. Orach, I. Chibwe, G. Bwire, O. C. Stine, A. M. Milstone, J. Lessler, A. S. Azman, W. Luo, K. Murt, D. A. Sack, A. K. Debes, S. Wohl, New Vibrio cholerae sequences from Eastern and Southern Africa alter our understanding of regional cholera transmission. medRxiv, doi: 10.1101/2024.03.28.24302717 (2024).

14. E. Ekeng, S. Tchatchouang, B. Akenji, B. B. Issaka, I. Akintayo, C. Chukwu, I. D. Dano, S. Melingui, S. Ousmane, M. O. Popoola, A. Nzouankeu, Y. Boum, F. Luquero, A. Ahumibe, D. Naidoo, A. Azman, J. Lessler, S. Wohl, Regional sequencing collaboration reveals persistence of the T12 Vibrio cholerae O1 lineage in West Africa. Elife 10 (2021).

15. G. Mboowa, N. L. Matteson, C. K. Tanui, M. Kasonde, G. K. Kamwiziku, O. A. Akanbi, J. J. E. Chitio, M. Kagoli, R. G. Essomba, A. Ayitewala, I. Ssewanyana, V. J. Ngo Bitoungui, A. A. Amuri, S. Azman, O. A. Babatunde, B. M. Akenji, A. Broban, E. B. Malembaka, F. Ongole, C. E. Chukwu, N. Ismael, O. Kapona, O. Laurindo, P. K. Mbala, G. A. Etoundi Mballa, I. C. Z. Miambo, A. A. Mwanyongo, G. Najjuka, J. Mutale, K. Musonda, A. M. Niyonzima, M. E. Nyenje, M. Popoola, D. M. Shempela, C. Medi Sike, S. M. Sitoe, D. W. Wanjohi, P. O. Welo, M. Yelewa, S. Yennan, L. Ziba, CholGEN Consortium, J. E. Bitilinyu-Bangoh, R. Chilengi, H. Hadja, J. Idris, J. P. M. Langa, D. Mukadi-Bamuleka, S. Nabadda, A. K. Debes, D. A. Sack, J. Kaseya, Y. K. Tebeje, S. Wohl, S. K. Tessema, Multicountry genomic analysis underscores regional cholera spread in Africa, medRxiv (2024) p. 2024.11.15.24317392.

16. A. Mutreja, D. W. Kim, N. R. Thomson, T. R. Connor, J. H. Lee, S. Kariuki, N. J. Croucher, S. Y. Choi, S. R. Harris, M. Lebens, S. K. Niyogi, E. J. Kim, T. Ramamurthy, J. Chun, J. L. N. Wood, J. D. Clemens, C. Czerkinsky, G. B. Nair, J. Holmgren, J. Parkhill, G. Dougan, Evidence for several waves of global transmission in the seventh cholera pandemic. Nature 477, 462–465 (2011).

17. S. Moore, A. Z. Dongdem, D. Opare, P. Cottavoz, M. Fookes, A. Y. Sadji, E. Dzotsi, M. Dogbe, F. Jeddi, B. Bidjada, M. Piarroux, O. T. Valentin, C. K. Glèlè, S. Rebaudet, A. G. Sow, G. Constantin de Magny, L. Koivogui, J. Dunoyer, F. Bellet, E. Garnotel, N. Thomson, R. Piarroux, Dynamics of cholera epidemics from Benin to Mauritania. PLoS Negl. Trop. Dis. 12, e0006379 (2018).

18. V. D. Blondel, J.-L. Guillaume, R. Lambiotte, E. Lefebvre, Fast unfolding of communities in large networks. J. Stat. Mech. 2008, P10008 (2008).

19. A. Lancichinetti, S. Fortunato, Consensus clustering in complex networks. Sci. Rep. 2, 336 (2012).

20. M. D. Karcher, J. A. Palacios, T. Bedford, M. A. Suchard, V. N. Minin, Quantifying and mitigating the effect of preferential sampling on phylodynamic inference. PLoS Comput. Biol. 12, e1004789 (2016).

21. S. Guindon, N. De Maio, Accounting for spatial sampling patterns in Bayesian phylogeography. Proc. Natl. Acad. Sci. U. S. A. 118, e2105273118 (2021).

22. G. W. Hassler, A. Magee, Z. Zhang, G. Baele, P. Lemey, X. Ji, M. Fourment, M. A. Suchard, Data integration in Bayesian phylogenetics. Annu. Rev. Stat. Appl. 10, 353–377 (2023).

23. N. Taty, D. Bompangue, S. Moore, J. J. Muyembe, N. M. de Richemond, Spatiotemporal dynamics of cholera hotspots in the Democratic Republic of the Congo from 1973 to 2022. BMC Infect. Dis. 24, 360 (2024).

24. N. J. Croucher, A. J. Page, T. R. Connor, A. J. Delaney, J. A. Keane, S. D. Bentley, J. Parkhill, S. R. Harris, Rapid phylogenetic analysis of large samples of recombinant bacterial whole genome sequences using Gubbins. Nucleic Acids Res. 43, e15 (2015).

25. A. J. Page, B. Taylor, A. J. Delaney, J. Soares, T. Seemann, J. A. Keane, S. R. Harris, SNP-sites: rapid efficient extraction of SNPs from multi-FASTA alignments. Microb. Genom. 2, e000056 (2016).

26. B. Q. Minh, H. A. Schmidt, O. Chernomor, D. Schrempf, M. D. Woodhams, A. von Haeseler, R. Lanfear, IQ-TREE 2: New models and efficient methods for phylogenetic inference in the genomic era. Mol. Biol. Evol. 37, 1530–1534 (2020).

27. S. Tavaré, Some probabilistic and statistical problems in the analysis of DNA sequences. Lecture of mathematics for life science 7, 57 (1986).

28. P. Sagulenko, V. Puller, R. A. Neher, TreeTime: Maximum-likelihood phylodynamic analysis. Virus Evol. 4, vex042 (2018).

29. F. Lemoine, O. Gascuel, Gotree/Goalign: toolkit and Go API to facilitate the development of phylogenetic workflows. NAR Genom. Bioinform. 3, lqab075 (2021).

30. M. A. Suchard, P. Lemey, G. Baele, D. L. Ayres, A. J. Drummond, A. Rambaut, Bayesian phylogenetic and phylodynamic data integration using BEAST 1.10. Virus Evol. 4, vey016 (2018).

31. D. L. Ayres, M. P. Cummings, G. Baele, A. E. Darling, P. O. Lewis, D. L. Swofford, J. P. Huelsenbeck, P. Lemey, A. Rambaut, M. A. Suchard, BEAGLE 3: Improved performance, scaling, and usability for a high-performance computing library for statistical phylogenetics. Syst. Biol. 68, 1052–1061 (2019).

32. A. Rambaut, A. J. Drummond, D. Xie, G. Baele, M. A. Suchard, Posterior summarization in Bayesian phylogenetics using tracer 1.7. Syst. Biol. 67, 901–904 (2018).

33. T. Bedford, S. Riley, I. G. Barr, S. Broor, M. Chadha, N. J. Cox, R. S. Daniels, C. P. Gunasekaran, A. C. Hurt, A. Kelso, A. Klimov, N. S. Lewis, X. Li, J. W. McCauley, T. Odagiri, V. Potdar, A. Rambaut, Y. Shu, E. Skepner, D. J. Smith, M. A. Suchard, M. Tashiro, D. Wang, X. Xu, P. Lemey, C. A. Russell, Global circulation patterns of seasonal influenza viruses vary with antigenic drift. Nature 523, 217–220 (2015).

34. P. Lemey, A. Rambaut, T. Bedford, N. Faria, F. Bielejec, G. Baele, C. A. Russell, D. J. Smith, O. G. Pybus, D. Brockmann, M. A. Suchard, Unifying viral genetics and human transportation data to predict the global transmission dynamics of human influenza H3N2. PLoS Pathog. 10, e1003932 (2014).

35. P. Lemey, A. Rambaut, A. J. Drummond, M. A. Suchard, Bayesian phylogeography finds its roots. PLoS Comput. Biol. 5, e1000520 (2009).

36. D. R. McDonald, On the Poisson Approximation to the Multinomial Distribution. The Canadian Journal of Statistics / La Revue Canadienne de Statistique 8, 115–118 (1980).

37. L. Damiano, B. Peterson, M. Weylandt, A tutorial on hidden Markov models using Stan. [Preprint] (2017). https://luisdamiano.github.io/stancon18/hmm_stan_tutorial.pdf.

38. C. Viboud, O. N. Bjørnstad, D. L. Smith, L. Simonsen, M. A. Miller, B. T. Grenfell, Synchrony, waves, and spatial hierarchies in the spread of influenza. Science 312, 447–451 (2006).

39. A. R. Tuite, J. Tien, M. Eisenberg, D. J. D. Earn, J. Ma, D. N. Fisman, Cholera epidemic in Haiti, 2010: using a transmission model to explain spatial spread of disease and identify optimal control interventions. Ann. Intern. Med. 154, 593–601 (2011).

40. G. D. Forney, The viterbi algorithm. Proc. IEEE Inst. Electr. Electron. Eng. 61, 268–278 (1973).

41. R Core Team, R: A Language and Environment for Statistical Computing. R Foundation for Statistical Computing [Preprint] (2024). https://www.R-project.org/.

42. Stan Development Team, Stan Reference Manual (2024; https://mc-stan.org).

43. J. Gabry, R. Češnovar, A. Johnson, cmdstanr: R Interface to “CmdStan.” [Preprint] (2024). https://mc-stan.org/cmdstanr/.

