## Supplementary Materials for "Defining Epidemiologically Relevant Units of Cholera Transmission in sub-Saharan Africa"

**This file contains:**

Materials and Methods
Figs. S1 to S7
Tables S2 to S4
References

**Other supplementary materials for this manuscript include**

Table S1 (csv)

### Materials and Methods

#### Data Sources

Sequence data were compiled from published sequences from GenBank and VibrioWatch from 1970 to 2023 (**table S1**). This curated list of cholera sequence data also contained metadata on country, year, and lineage call. Cholera case data for this analysis came from the WHO's database of cholera cases reported by Member States, which contains annual reported suspected and confirmed cholera cases by country from 1970 to 2023.

#### Phylogeographic analysis

We used the phylogeny subcommand of bacpage (Bacterial Genomic Analysis Pipeline; github.com/CholGen/bacpage) to generate a recombination-masked maximum likelihood phylogeny. Briefly, the pipeline concatenated individual reference-based assemblies along with the N16961 reference into a pseudo-alignment. Problematic sites including known recombinant regions, repetitive sequences, and homoplastic sites were masked based on the GFF file provided by Weill et al. 2017 (*12*). Novel recombinant regions and regions with significantly elevated substitution densities were masked using Gubbins v2.3.4 (*24*). To reduce the computational burden of phylogenetic analysis, we filtered the pseudo-alignment to include only variable positions using SNP-sites v2.5.1 (*25*). We then constructed a maximum likelihood phylogenetic tree using IQ-TREE2 (*26*) with a GTR substitution model (*27*).

The resulting phylogeny was rooted on the reference genome. To further reduce computational burden, we extracted the subtree descending from the most recent common ancestor of lineages AFR9-15, corresponding to the third wave of 7PET. This subtree was time-resolved using Treetime v0.11.3 (*28*). We pruned taxa that deviated more than three interquartile ranges from the clock-rate regression from the phylogeny. Additionally, we randomly resolved polytomies by adding zero-length branches with gotree v0.4.5 (*29*).

We refined the time-resolved phylogeny using BEAST X v10.5.0 (beta 5) (*30*). We specified a GTR substitution model with gamma-distributed rate heterogeneity under an uncorrelated relaxed molecular clock and a constant coalescent tree prior. For the uncorrelated relaxed molecular clock, we specified an informative prior consisting of a log-normal distribution with a mean of $8.4\times10^{-7}$ substitutions/site/year and a standard deviation of $1.4\times10^{-6}$, consistent with parameters reported by Weill et al. 2017 (*12*).

We ran a Markov Chain Monte Carlo (MCMC) chain of 300 million steps utilizing the BEAGLE computational library (*31*), sampling parameters and trees every 10,000 and 100,000 states, respectively. We assessed the convergence and mixing of the MCMC chain with Tracer v1.7.2 (*32*), and confirmed that all estimated continuous parameters achieve effective sample sizes of greater than 100.

We performed a discrete state ancestral reconstruction on geographic states using BEAST. This analysis reconstructed location-transition history across an empirical distribution of 2000 time-calibrated trees sampled from the posterior tree distribution estimated above (*33*, *34*). For this analysis, we assigned each taxon a discrete location state based on its country of collection. To limit the number of transition rates requiring estimation, we binned non-African sequences into a single "Other" state. We assumed geographic transition rates were reversible and used a symmetric substitution model. We employed Bayesian stochastic search variable selection to infer non-zero migration rates (*35*). The MCMC algorithm ran for 500,000 generations and transition rates were sampled every 500 generations. We used TreeAnnotator v1.10 to construct a maximum clade credibility (MCC) tree, which we visualized using Baltic (https://github.com/evogytis/baltic).

Modeling presence and prevalence of cholera lineages (HMM Model)

We modeled the occurrence of distinct cholera lineages in countries in sub-Saharan Africa through time using a Hidden Markov Model (HMM). Our targets of interest were (1) the strength of connectivity driving cholera transmission between locations, and (2) the underlying presence and prevalence of cholera lineages in each country across time.

We considered discrete, hidden states, $z_{v,i,t} \in \{0,1\}$, representing absence or presence of a cholera lineage $(v)$ in a given country $(i)$ and year $(t)$, focusing on the probability of lineage presence, $\Pr(z_{v,i,t} = 1)$ (see **table S4** for list of parameters and interpretations).

For modeling purposes, we made the following assumptions: (1) no competition between lineages, (2) introduction of a lineage into a given country is independent of other lineages already circulating in that country, (3) the probability of a lineage being introduced or re-introduced from another country is a function of the number of reported cholera cases in that country and on the distance separating the two countries, (4) perfect specificity of sequencing $[\Pr(y_{v,i,t} \geq 1 \mid z_{v,i,t} = 0) = 0]$, and (5) introduction of lineage $v$ to sub-Saharan Africa is based on inferred introduction timing and initial locations of circulation. Additionally, due to complexities in disentangling and modeling cases/lineages in Sudan & South Sudan before and after 2014, we geographically combined them in analyses, fully recognizing that the two countries are politically separate and may have different transmission dynamics.

We constructed our model, $\boldsymbol{H} = \{\pi, \boldsymbol{\Phi}, \boldsymbol{\Psi}\}$, as follows:

$$\pi_{v,i,1}\text{: initial probability of presence, } \Pr\left(z_{v,i,1} = 1\right)$$
$$\boldsymbol{\Phi}\text{: transition process, } \Phi_{k,l} = \Pr\left(z_{v,i,t} = 1 \,\middle|\, z_{v,1,t-1}, \dots, z_{v,M,t-1}\right)$$
$$\boldsymbol{\Psi}\text{: observation process, } \psi_l = \Pr\left(y_{v,i,t} \middle| z_{v,i,t}\right)$$

*Initial Probability of Presence*

The initial probability of presence, $\pi_{v,i,1}$, was inferred based on lineage observation, using priors assuming (1) high probability of presence if the lineage was observed in the country of interest, (2) moderate probability if observed elsewhere, (3) moderately low probability if inferred to have been introduced to the continent before 1970, and (4) low probability if not observed on the continent and not inferred to have already been introduced to the continent (see *Parameter Priors* below).

*Observation Process*

The observed data are the number of sequenced samples of lineage $v$ in country $i$ and year $t$, $y_{v,i,t}$, and reported suspected cholera cases in country $i$ and year $t$, $c_{i,t}$.

Sequence Data:

The observation process for sequence data, $\boldsymbol{\Psi}$, is $\boldsymbol{Y}_{v,i} = [Y_{v,i,t}, t = 1, \dots, T]$, is the observation of the number of sequenced samples of lineage $v$ in country $i$ and year $t$, and is associated with the hidden process, $\boldsymbol{\Phi} = \left(\Phi^{k,l}_{v,i,t}, t = 1, \dots, T\right)$, of the underlying true presence of lineage $v$ in country $i$ and year $t$ (see *Transition Process*, below). To estimate lineage prevalence, we modeled the probability of our observed sequence data $(\boldsymbol{y}_t)$ given the unobserved (hidden) states of presence $(z_{v,i,t} = 1)$ or absence $(z_{v,i,t} = 0)$ of lineage $v$ in country $i$ at time $t$:

$$\Pr(y_{v,i,t}|z_{v,i,t}) = \begin{cases} (1-\rho_{v,i,t}) + \rho_{v,i,t}\Pr(y_{v,i,t}=0|\lambda_{v,i,t}) & \text{if } y_{v,i,t}=0 \\ \rho_{v,i,t}\Pr(y_{v,i,t}|\lambda_{v,i,t}) & \text{if } y_{v,i,t}>0 \end{cases}$$

where $\rho_{v,i,t} = \Pr(z_{v,i,t} = 1)$ and $\lambda_{v,i,t}$ is the number of sequences attributable to lineage $v$ in country $i$ and year $t$. We use the Poisson approximation of the multinomial (*36*) in our observation process for sequenced cholera lineages:

$$Y_{v,i,t} \sim Poisson(\lambda_{v,i,t})$$

$$\log(\lambda_{v,i,t}) \sim Normal\left(\log\left[\frac{I(c_{i,t}>0)\,\overline{\boldsymbol{y_{i,t}}}\,E(z_{v,i,t})}{E(\sum_{q\neq v} z_{q,i,t}|z_{v,i,t}=1)}\right], \sigma\right)$$

$$\lambda^*_{v,i,t} = \frac{\lambda_{v,i,t}}{\sum_v \lambda_{v,i,t}}$$

Reported Suspected Cholera Cases:

We also modeled the observation process for cholera cases ($c_{i,t}$):

$$\Pr(c_{i,t}|z_{v,i,t}) = \begin{cases} (1-\alpha^+_{i,t}) + \alpha^+_{i,t}\Pr\left(c_{i,t}=0\middle|\frac{1}{\varepsilon_i}+N_{i,t}\right) & \text{if } c_{i,t}=0 \\ \alpha^+_{i,t}\Pr\left(c_{i,t}\middle|\frac{c_{i,t}}{\varepsilon_i}\right) & \text{if } c_{i,t}>0 \end{cases}$$

Where $\varepsilon_i$ is case underreporting by country and $N_{i,t}$ is the total number of sequences collected by country and year. Given that there are a small number of instances where we observe sequences in countries and years with no reported cases, we handle these instances by assuming there are at least as many cases as there are observed sequences, allowing for underreporting. Additionally, $\alpha^+_{i,t}$ is the probability of any lineage being present, based on the forward algorithm:

$$\alpha^+_{i,t} = 1 - \prod_v 1 - \alpha_{v,i,t}$$

*Transition Process*

We modeled the transition process between presence states as a function of within-country persistence, extra-continental introductions, and estimated transnational connectivity.

Specifically, for the transition process, $\boldsymbol{\Phi}$, lineage presence is based on the probability of establishment or persistence of lineage $v$ in country $i$ at time $t$. The probability of presence in time $t$ given the state in time $t-1$, is based on the transition matrix $\mathbf{A} = \{\Phi^{k,l}_{v,i,t}\}$, where $\Phi^{k,l}_{v,i,t} = \Pr(z_{v,i,t} = 1\,|z_{v,1,t-1}, \ldots, z_{v,M,t-1})$, is a function of the introduction/re-introduction rate, $\phi_{v,j,t}$ from other countries. Because we focus on the probability of lineage presence, we define $\Phi_{v,i,t} = \Pr(z_{v,i,t} = 1\,|\,z_{v,i,t-1} = k)$:

$$\Phi_{v,i,t}^{k,l} = 1 - \left[ \left(1 - \gamma_{v,i,t}\right) \prod_j 1 - \left(1 - e^{-\phi_{v,j,t}}\right) \right]$$

where:

$$1 - e^{-\phi_{v,j,t}} = \Pr\left(z_{v,i,t} = 1 | z_{v,j,t-1} = k\right)$$

$$\phi_{v,j,t} = \left(\lambda_{v,j,t-1}^{*} c_{j,t-1}\right)^{\eta} \xi_{j,i}$$

Where introduction to country *i* is based on introduction from outside of the continent, $\gamma_{v,i,t}$, and introduction/reintroduction from within the continent (cross-border transmission or within-country persistence). Persistence and transnational transmission are a function of the number of cholera cases attributable to lineage v, calculated from prevalence of lineage $v$ $(\lambda_{v,j,t-1}^{*})$ and total observed cases $c_{j,t-1}$, in country *j* and year *t-1*, and connectivity between locations. Prevalence is calculated relative to other circulating lineages:

$$\lambda_{v,j,t-1}^{*} = \frac{\lambda_{v,j,t-1}\alpha_{v,j,t-1}}{\sum_v \lambda_{v,j,t-1}\, \alpha_{v,j,t-1}}$$

where:

$$\alpha_{v,j,t-1} = \Pr\left(z_{v,j,t-1} = 1 | z_{v,j,t-2} = k\right)$$

Connectivity between locations, $\xi_{j,i}$, is modeled using a gravity model for locations $j \neq i$, otherwise is the self-persistence parameter, $\delta$:

$$\log\left(\xi_{j,i}\right) = \begin{cases} \log(\delta) & \text{if } j = i \\ \log\left(\kappa \dfrac{pop_j^{\tau_d} pop_i^{\tau_r}}{d_{j,i}^{\zeta}}\right) + \omega_{j,i} & \text{if } j \neq i \end{cases}$$

Where $\kappa$ is the gravity constant, $pop_i$ is the population size in country $i$, $d_{j,i}$ is the Euclidean distance between centroids of countries i and j in kilometers, $\omega_{j,i}$ represents the random effect, $\tau_d$ and $\tau_r$ are scaling factors for population size for donor and recipient countries, respectively, and $\zeta$ is a scaling factor for distance between country centroids.

We fit the spatial random effect using a mixture model designed to force the model to assume locations were disconnected unless there was evidence of connection, where $\theta_k$=0.7, $\mu_k$= [-3, 0], $\sigma_k$ = [1,1] based on functional form analysis:

$$\Pr\left(\omega_{j,i} | \theta_k, \mu_k, \sigma_k\right) = \sum_k \theta_k Normal(\mu_k, \sigma_k)$$

Finally, intercontinental introduction, $\gamma_{v,i,t}$, was calculated inferred introduction timings ($\gamma_{v,t}$) and locations ($\gamma_{v,i}$) of lineages:

$$\gamma_{v,i,t} = \gamma_{v,t} \times \gamma_{v,i}$$

*Forward, backward, and forward-backward algorithm*

With the elements of the HMM as defined above, we then implemented the forward algorithm to calculate filtered probabilities of presence, $\Pr(z_{v,i,t} = 1 \mid \boldsymbol{y}_{1:t})$ (*37*):

$$\Pr(z_{v,i,t} = 1 \mid \boldsymbol{y}_{1:t-1}) = \sum_{k \in \{0,1\}} \Phi_{v,i,t}^{k,l} \Pr(z_{v,i,t} = 1 \mid \boldsymbol{y}_{1:t-1})$$

$$\psi_{v,i,t} = \Pr(y_{v,i,t} | z_{v,i,t})$$

$$\alpha_{v,i,t}^{l} = \alpha_{v,i,t-1}^{k} \Phi_{v,i,t}^{k,l} \psi_{v,i,t}$$

$$\alpha_{v,i,t} = \Pr(z_{v,i,t} = 1 | z_{v,i,t-1} = k)$$

Where $\alpha_{v,i,t}$ is the forward probability of presence, $\Phi_{v,i,t}^{k,l}$ is the transition probability (i.e., probability of introduction/re-introduction into country $i$ at time $t$), and $\psi_{v,i,t}$ is the observation process.

The backward algorithm was implemented as:

$$\beta_{v,i,t}(l) = \Pr(y_{v,i,t+1:T} | z_{v,i,t} = l)$$

$$\beta_{v,i,t-1}(k) = \Pr(y_{v,i,t:T} | z_{v,i,t} = k)$$

$$= \sum_{l=1}^{L} \Pr(y_{v,i,t+1:T} | z_{v,i,t} = l) \Pr(\boldsymbol{y}_t | z_{v,i,t} = l) \Pr(z_{v,i,t} = l | z_{v,i,t-1} = k)$$

$$= \beta_{v,i,t} \psi_{v,i,t} \Phi_{v,i,t}^{k,l}$$

With the backward algorithm specified, we define the forward-backward algorithm for the smoothed probability of presence/absence of a specific lineage in a given country and year ($\rho_{v,i,t}^{k}$) as:

$$\rho_{v,i,t}^{k} = \Pr(z_{v,i,t} = l \mid \boldsymbol{y}_{1:T})$$

$$= \frac{\alpha_{v,i,t}(l) \beta_{v,i,t}(l)}{\Pr(\boldsymbol{y}_{1:T})}$$

$$\approx \alpha_{v,i,t}(l) \beta_{v,i,t}(l)$$

*Parameter Priors*

Initial Probability:

$$\pi_{v,i,1} \sim \begin{cases} Beta(a_{\pi_1}, b_{\pi_1}): E(\pi_1) = 0.98, \sigma_{\pi_1} = 0.02, & y_{v,i,t} > 0 \\ Beta(a_{\pi_2}, b_{\pi_2}): E(\pi_2) = 0.43, \sigma_{\pi_2} = 0.18, & y_{v,t} > 0 \\ Beta(a_{\pi_3}, b_{\pi_3}): E(\pi_3) = 0.17, \sigma_{\pi_3} = 0.10, & y_{v,t-} > 0 \\ Beta(a_{\pi_4}, b_{\pi_4}): E(\pi_4) = 0.02, \sigma_{\pi_4} = 0.02, & y_{v,t-} = 0 \end{cases}$$

Observation Process:

Case underreporting by country was fit based on the following prior distribution:

$$\varepsilon_i = Beta(a_\varepsilon, b_{\varepsilon)}: E(\varepsilon) = 0.8, \sigma_\varepsilon = 0.1$$

Transition Process:

The connectivity parameters were fit based on the following prior distributions:

$$\eta \sim Beta(a_\eta, b_\eta): E(\eta) = 0.45, \sigma_\eta = 0.002$$
$$\kappa \sim Normal(0.5, 0.1)$$
$$\log(\delta) \sim Normal(\text{-}3, 0.5)$$
$$\tau_d \sim Beta(a_{\tau_d}, b_{\tau_d}): E(\tau_d) = 0.45, \sigma_{\tau_d} = 0.002$$
$$\tau_r \sim Beta(a_{\tau_r}, b_{\tau_r}): E(\tau_r) = 0.35, \sigma_{\tau_r} = 0.002$$
$$\zeta \sim Normal(2.25, 0.2)$$

Priors were chosen based on functional form analyses, with gravity priors informed by literature estimates of gravity parameters for epidemic spread (*38*), including cholera transmission (*39*). Tight priors were required for case and population transformation parameters for model convergence. Priors for lineage introduction timings ($\gamma_{v,t}$) and locations ($\gamma_{v,i}$) from outside of the continent were based on time to most recent common ancestor (TMRCA) findings from Weill et al. 2017 for lineages AFR1-AFR8 (*12*), our phylogeographic analysis for lineages AFR9-AFR15 (**table S5**), and Xiao et al. 2025 for lineages AFR16-AFR17 (*13*).

*Model Diagnostics*

The HMM model was run using 4 chains with 2000 warmup and 2000 sampling iterations. Chain convergence was assessed by whether any divergent transitions occurred, visual inspections of trace plots and posterior parameter distributions, R-hat values (≤1.1), and ESS.

*Post-estimation*

We filled in the gaps in the observed phylogenetic lineage data by inferring the prevalence of each lineage across space and time and inferring the most likely sequence of (presence) states for each lineage using the Viterbi algorithm (*40*).

First, using smoothed predicted probabilities of presence and predicted lineage prevalence, we inferred the likely number of cases attributed to each lineage across space and time based on a multinomial draw.

$$c^*_{v,i,t} = multinomial(V, \boldsymbol{\lambda}^*_{i,t})$$

And determined lineage-specific prevalence as:

$$\lambda^{\rho^+}_{v,i,t} = \lambda^*_{v,i,t}\boldsymbol{\rho}^+_{i,t}$$

where:

$$\boldsymbol{\rho}^+_{i,t} = 1 - \prod_v 1 - \rho_{v,i,t}$$

We then used the Viterbi algorithm (*37*, *40*) to infer the most likely sequence of states for each lineage across space and time. This algorithm uses maximum a posteriori estimation to jointly infer the single most likely state sequence, where the most probable path is calculated moving forward in time, conditioning on the most likely state determined for the previous time step to calculate the most likely state at time *t*.

$$\boldsymbol{z}^*_{i,t} = \text{argmax}_{\boldsymbol{z}_{v,i,1:T}} \Pr\left(\boldsymbol{z}_{v,i,1:T} \mid \boldsymbol{y}_{v,i,1:T}\right)$$

Defining ‘cholera transmission units’

To define epidemiologically relevant ‘cholera transmission units,’ we used transition rates from the full posterior of the phylogeographic analysis and inferred transnational connectivity from the full posterior from the HMM model. For each analysis, we used the Louvain algorithm for community detection (*18*) to define non-overlapping communities on the full network, using transition rates and inferred connectivity as edge weights for the phylogeographic and HMM analyses, respectively. This method of community detection produces a distribution of the proportion of times a country-pair is assigned to the same community (**Fig. S1**). We then used a consensus clustering algorithm (*19*) to define cholera transmission units for both phylogeographic and HMM analyses. Briefly, to implement this algorithm, we use the proportion of times a country-pair is assigned to the same community as edge weights, with a threshold $\tau$ of 0.15 for edge weights. Any edge weight below the threshold is assigned a weight of 0, and the algorithm is run until there are only 2 possible edge weights (0 or 1), with each country now assigned to a consensus cluster. We confirmed the assignment of cholera transmission units using a divisive clustering algorithm (DIANA) where we defined the dissimilarity matrix based on the output of the proportion of times a country-pair clustered together from the Louvain algorithm, with the number of clusters (k) chosen based on the silhouette method, finding identical consensus clusters between the two methods.

Simulation of cholera spread

Using inferences from our HMM model, we then conducted simulations to predict dynamics of cholera spread based on introductions or re-introductions of a cholera lineage into each country.

We first simulated the one-year risk of an outbreak in countries across the continent based on an introduction or re-emergence in each country. This simulation was set up based on the transition process from our HMM model, defined above. Briefly, we defined the probability of an outbreak $p_{i,t=1}$ in each country $i$ in year $t = 1$ based on the number of cases in the country with an outbreak (source country, $j$) in year $t - 1$ and transnational connectivity country $\xi_{i,j}$:

$$p_{i,t} = binomial(1, \alpha_{i,t})$$

$$\alpha_{i,t} = 1 - \Phi_{i,t}$$

$$\Phi_{i,t} = 1 - \left(1 - e^{-\phi_{i,t}}\right)$$

where:

$$1 - e^{-\phi_{i,t}} = \Pr\left(z_{i,t} = 1 | z_{j,t-1} = k\right)$$

$$\phi_{i,t} = \left(c_{j,t-1}\right)^{\eta} \xi_{j,i}$$

The cases in the source country ($j$) were sampled from a Poisson distribution based on the observed distribution of reported suspected cholera cases in country $j$ from 1990 onwards, due to sparse reporting in early years.

We then examined the downstream impacts of an outbreak due to a new or re-introduction of a lineage in country $i$, simulating the speed of spread across the continent. This simulation was conducted similarly to the previous simulation, except that the full network of transnational connectivity was taken into account and the simulation was run over 30 years:

$$p_{i,t} = binomial(1, \alpha_{i,t})$$

$$\alpha_{i,t} = 1 - \Phi_{i,t}$$

$$\Phi_{i,t} = \prod_{j} 1 - \left(1 - e^{-\phi_{i,t}}\right)$$

where:

$$1 - e^{-\phi_{i,t}} = \Pr\left(z_{i,t} = 1 | z_{j,t-1} = k\right)$$

$$\phi_{i,t} = \left(c_{j,t-1}\right)^{\eta} \xi_{j,i}$$

Where again, cases were sampled from a Poisson distribution based on the distribution of cases observed in country $j$ from 1990 onwards.

The output from this simulation contained the distribution of the timing of arrival of a cholera lineage in each country $i$ given an outbreak or introduction/re-introduction in country $j$.

We ran 2000 simulations for each country of introduction (44 countries) for each scenario. Parameters $\xi_{j,i}, \eta,$ and $\delta$ were sampled from the full posterior of the HMM model.

Sensitivity analyses

First, cholera transmission dynamics likely evolve through time as risk factors change. Therefore, we subset the data to observations from 1990 onwards and from 2000 onwards and re-ran the HMM model and clustering analysis outlined above.

Next, to evaluate our model's ability to recover missing lineage information, we created multiple partitions where we downsampled outbreaks with sequence availability by 20%, meaning we removed information about detected lineages from 20% of reported outbreaks. We then re-ran the HMM model on each partition and determined the number of false negatives (known lineage presence, but not recovered by the model), false positives assuming clonal outbreaks (a lineage different from the observed detected lineage was inferred by the model in addition to the originally detected lineage), and false positives assuming nonclonal outbreaks (allowing inferred lineages in addition to lineage(s) detected through sequencing in observed data). We then calculated mean recall and precision assuming (1) clonal outbreaks and (2) nonclonal outbreaks across partitions (**table S2**).

Finally, to evaluate our predictions from speed of spread simulations, we compared simulated arrival times to observed arrival times. We determined observed arrival times as the time between estimated median (and bounds of 95% HPD) year of arrival into a likely country of introduction, based on findings from Weill et al. 2017 for early lineages, and our phylogeographic analysis for later lineages, and (1) the first year a lineage was detected in countries where the lineage was observed or (2) the first year a lineage was inferred to be present based on our HMM model. We calculated agreement between observed/inferred arrival times and median simulated arrival times using the Pearson correlation coefficient.

Software

Data cleaning, post-estimation analyses, and figure generation were conducted in R v4.3.3 (*41*). We used Stan (*42*) to run the HMM models, using cmdstanr v0.7.1 (*43*) to interface with cmdstan v2.34.1.

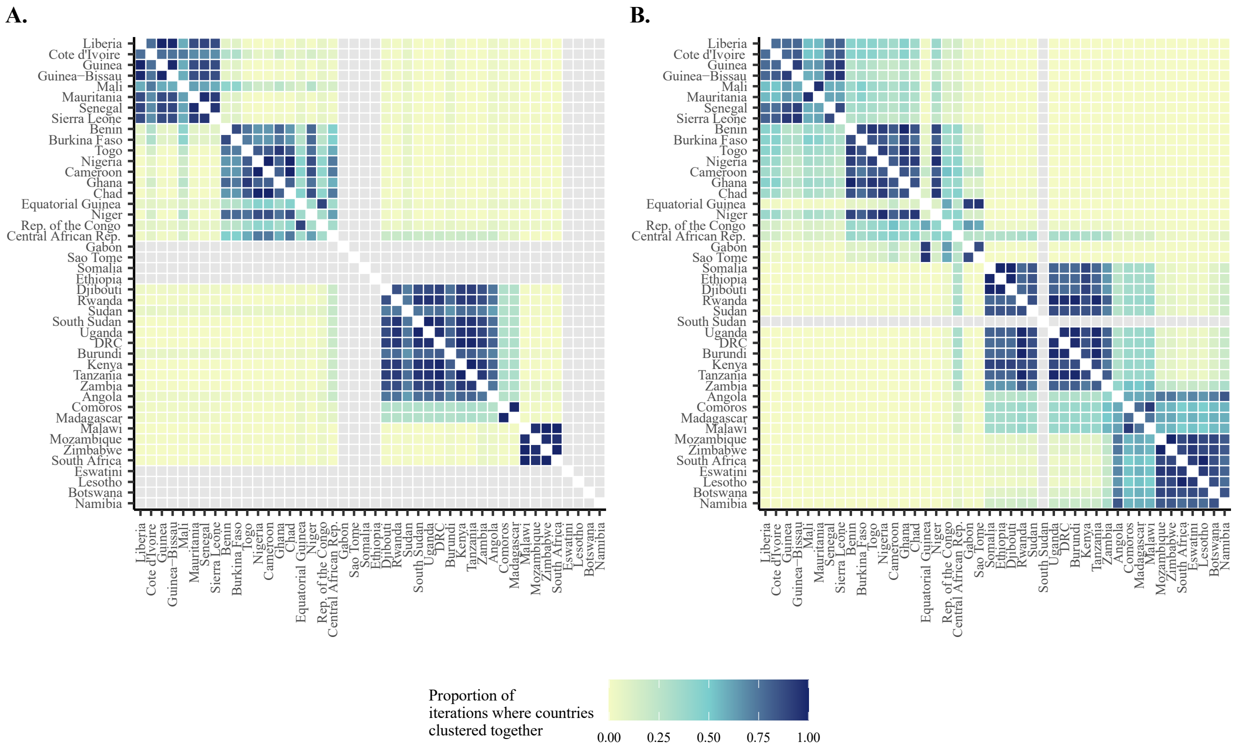


**Fig. S1. Proportion of iterations where country-pairs clustered together.** Heatmaps of the proportion of runs where country-pairs were grouped together in nonoverlapping neighborhoods using the Louvain method to define neighborhoods. (A) phylogeographic analysis, where an iteration refers to a state in the posterior, (B) HMM analysis.

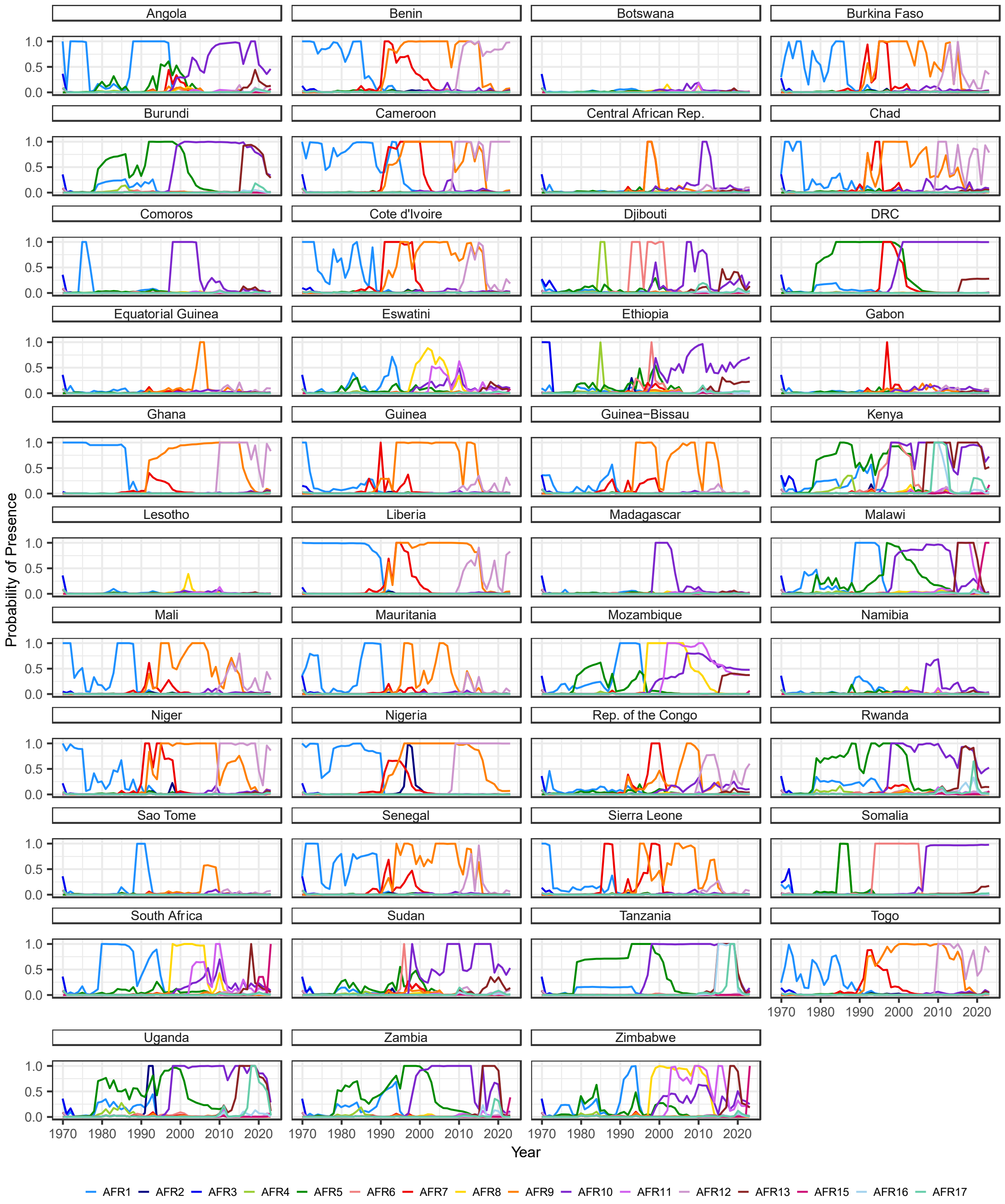


**Fig. S2. Inferred probability of presence of cholera lineages over time by country.** Facet panels show the mean probability of presence of each cholera lineage by year and country from the full posterior, with points in the top of each panel indicating whether a lineage was detected in that country and year.

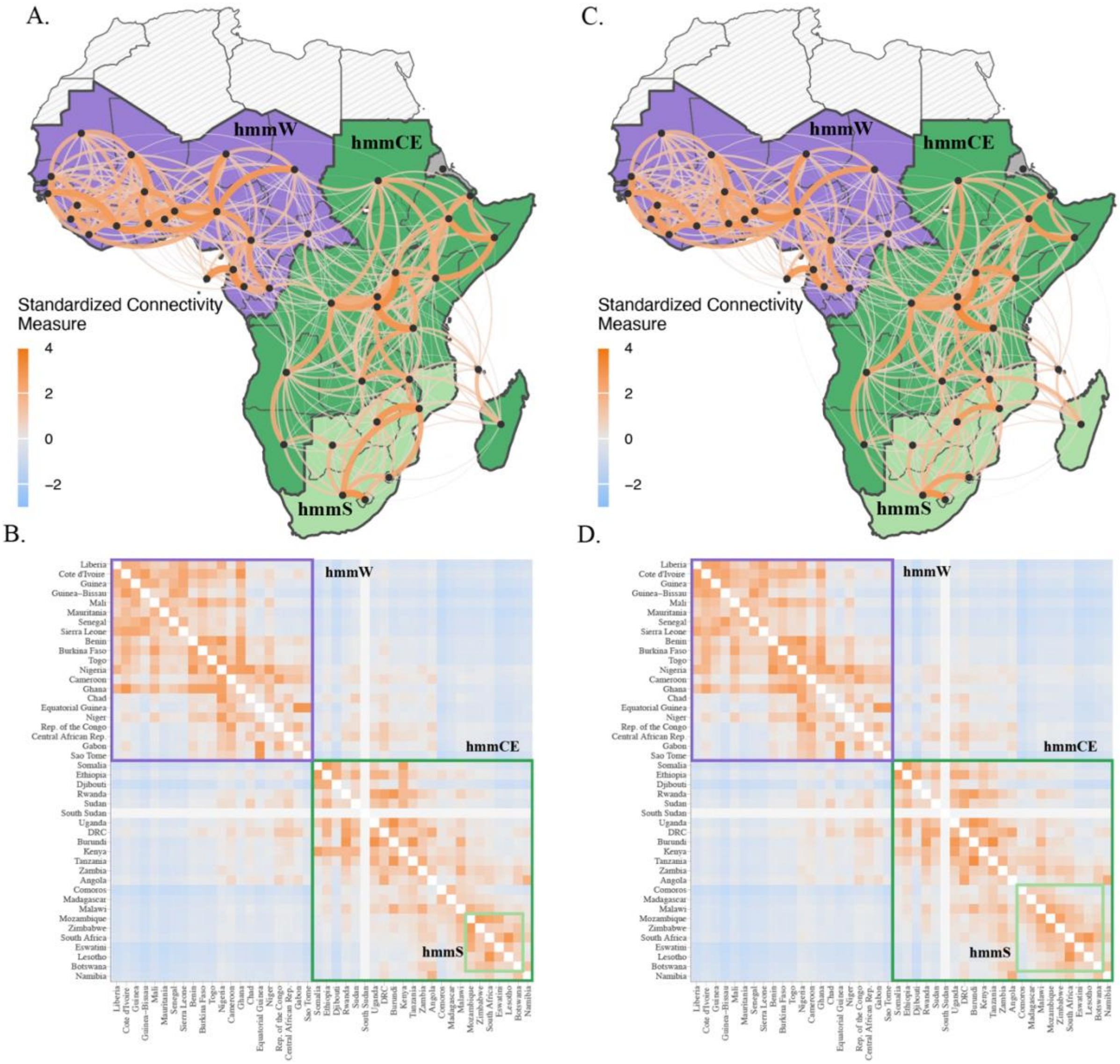


**Fig S3. Inferred cholera transmission units and cross-country transmission in sub-Saharan Africa. (A)** Results from HMM analysis where data was subsetted to 1990 onwards. Three identified cholera transmission units are shown by fill color (purple - hmmW, dark green - hmmCE, light green - hmmS), with the color and and thickness of edges representing the standardized connectivity measure between countries. Only the top 50% of edges are plotted. **(B)** heat map of the standardized connectivity measure between countries using the full network. **(C)** Results from HMM analysis where data was subsetted to 2000 onwards. Three identified cholera transmission units are shown by fill color (purple - hmmW, dark green - hmmCE, light green - hmmS), with the color and and thickness of edges representing the standardized connectivity measure between countries. Only the top 50% of edges are plotted. **(D)** heat map of the standardized connectivity measure between countries using the full network.

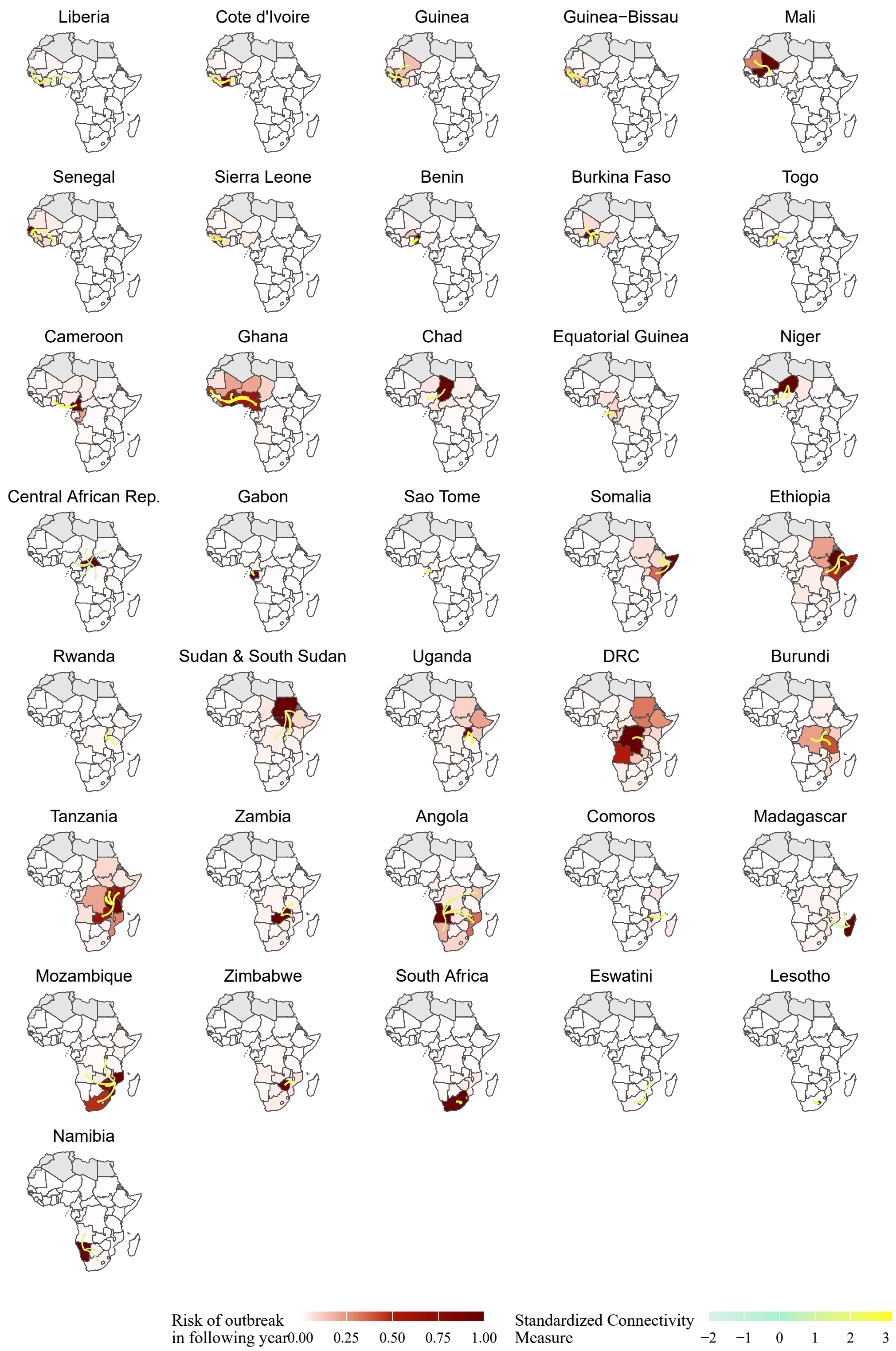


**Fig. S4. One-year risk of cholera outbreak by country of introduction.** Simulated one-year risk of cholera outbreaks from 2,000 introductions of cholera into each of 44 countries (sampling parameters from the posterior for each), with darker red indicating higher risk of an outbreak in the following year. Edges indicate the asymmetrical strength of connectivity between the country of introduction and the rest of the continent.

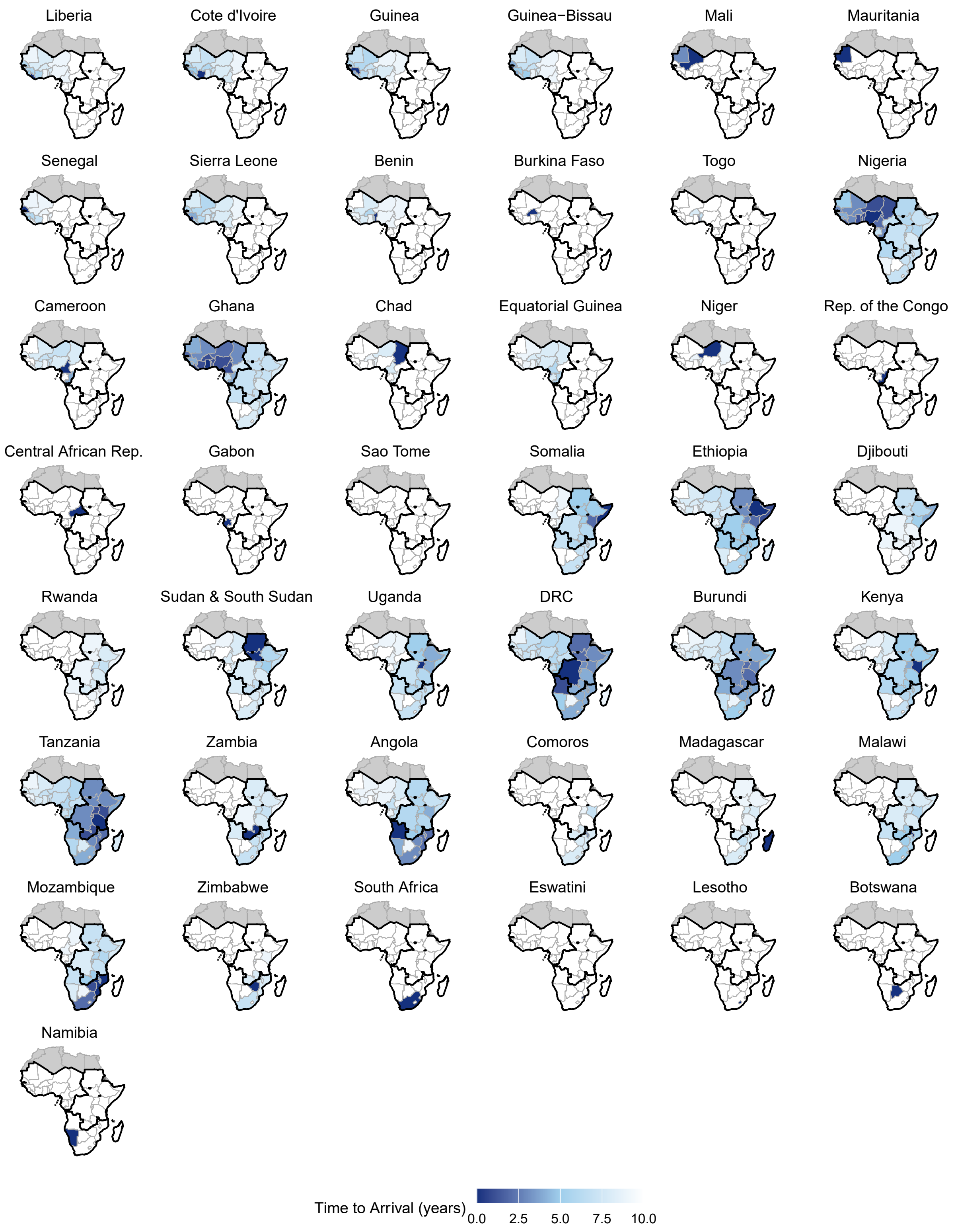


**Fig. S5. Speed of cholera spread by country of introduction.** Simulated downstream time to arrival of cholera from 2,000 introductions of cholera into each of 44 countries (sampling parameters from the posterior for each), with darker blue indicating higher risk of an outbreak in the following year.

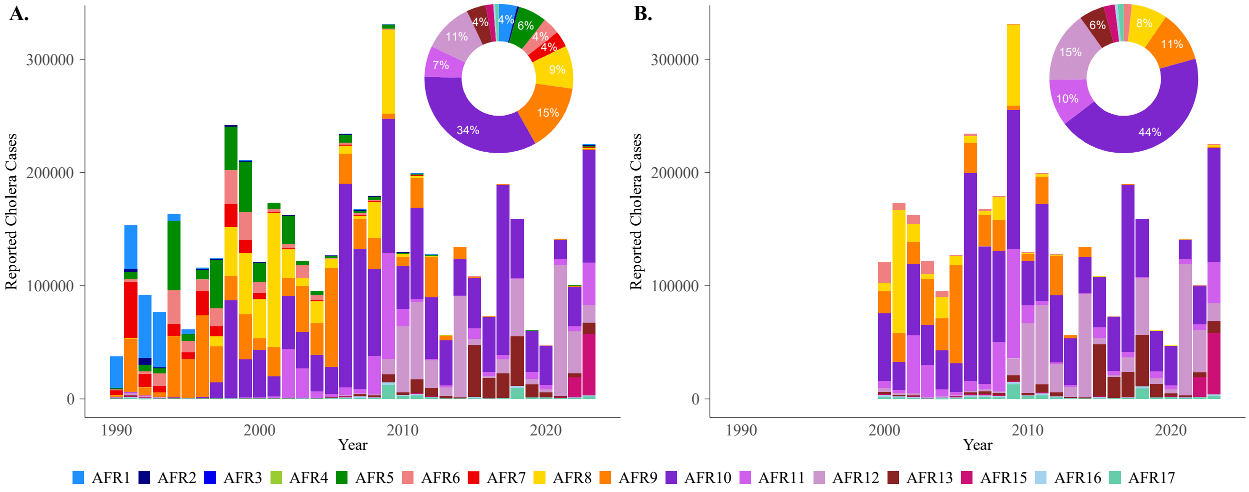


**Fig. S6. Lineage prevalence.** Bar graph indicates total cases by year attributable to a given observed lineage or lineage inferred from the HMM model, and donut plot shows the proportion of cases attributable to each lineage overall in **(A)** data subset to 1990 onwards, and **(B)** data subset to 2000 onwards.

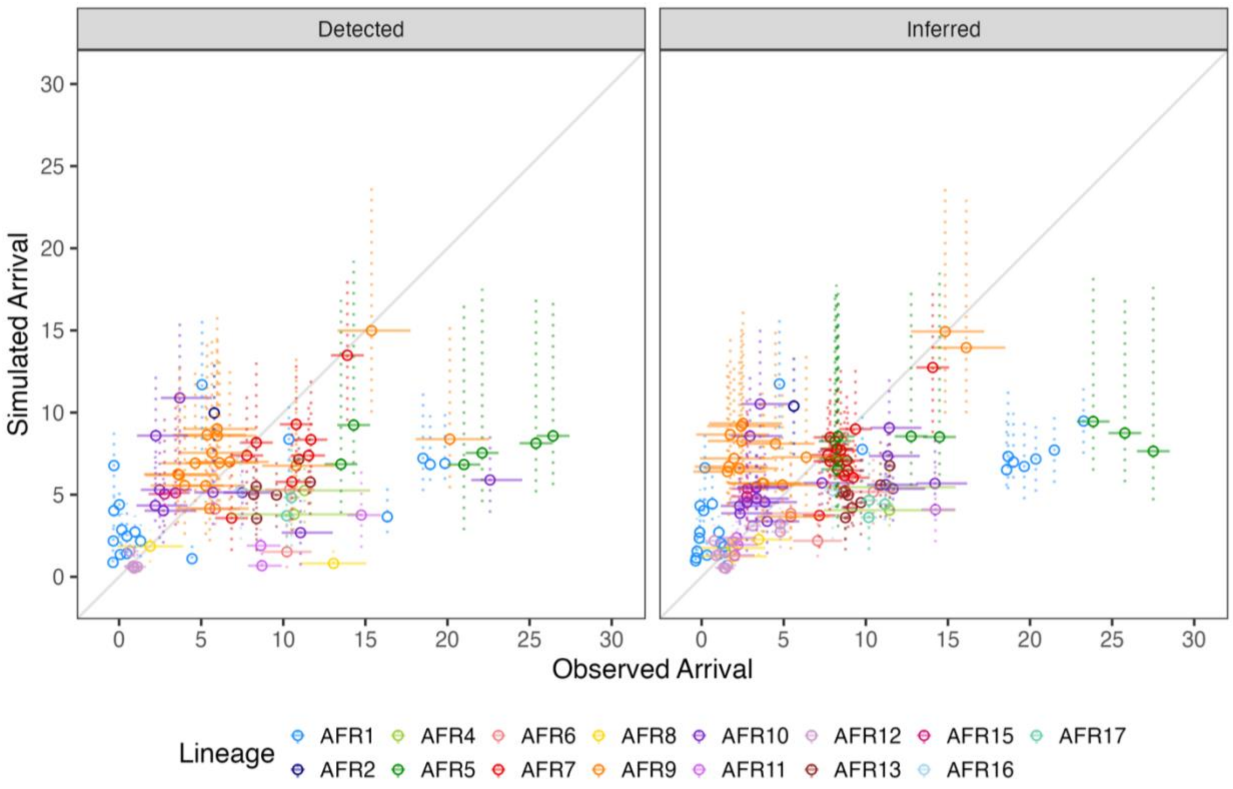


**Fig. S7. Observed vs simulated time to arrival by lineage.** Comparing median (IQR shown by dashed vertical line) simulated arrival times of lineages to **(left)** observed lineage arrival time, determined by inferred country and median (solid line depicts 95% HPD) year of introduction to the continent from phylogeographic analyses and first year the lineage was detected by country, and **(right)** inferred lineage arrival time, determined by inferred country and median (solid line depicts 95% HPD) year of introduction to the continent from phylogeographic analyses and first year the lineage was inferred by the HMM model to have arrived by country.

**Table S1 (separate .csv file).** List of accession numbers and metadata for sequences used in this analysis.

| Country with an Outbreak | Expected Outbreaks Caused in Additional countries in 1 Year | Outbreaks Caused Within vs. Between Transmission Units | | Outbreaks by Transmission Unit | | |
|---|---|---|---|---|---|---|
| | | Between Units | Within Unit | hmmW | hmmS | hmmCE |
| Angola | 1.50 | 0.55 | 0.95 | 0.18 | 0.95 | 0.37 |
| Benin | 0.49 | 0.02 | 0.47 | 0.47 | 0.01 | 0.01 |
| Botswana | 0.02 | 0.00 | 0.01 | 0.00 | 0.01 | 0.00 |
| Burkina Faso | 0.42 | 0.01 | 0.41 | 0.41 | 0.00 | 0.01 |
| Burundi | 1.51 | 0.22 | 1.29 | 0.03 | 0.20 | 1.29 |
| Cameroon | 0.73 | 0.11 | 0.63 | 0.63 | 0.04 | 0.07 |
| Central African Rep. | 0.09 | 0.04 | 0.05 | 0.05 | 0.00 | 0.04 |
| Chad | 0.42 | 0.12 | 0.30 | 0.30 | 0.02 | 0.10 |
| Comoros | 0.53 | 0.11 | 0.42 | 0.00 | 0.42 | 0.11 |
| Cote d'Ivoire | 0.84 | 0.02 | 0.83 | 0.83 | 0.01 | 0.01 |
| DRC | 2.64 | 1.14 | 1.50 | 0.41 | 0.73 | 1.50 |
| Djibouti | 0.36 | 0.03 | 0.33 | 0.02 | 0.01 | 0.33 |
| Equatorial Guinea | 0.85 | 0.06 | 0.79 | 0.79 | 0.02 | 0.04 |
| Eswatini | 0.06 | 0.01 | 0.05 | 0.00 | 0.05 | 0.01 |
| Ethiopia | 2.59 | 0.39 | 2.20 | 0.18 | 0.21 | 2.20 |
| Gabon | 0.21 | 0.02 | 0.19 | 0.19 | 0.01 | 0.01 |
| Ghana | 5.00 | 0.08 | 4.92 | 4.92 | 0.04 | 0.05 |
| Guinea | 0.88 | 0.02 | 0.86 | 0.86 | 0.01 | 0.01 |
| Guinea-Bissau | 0.78 | 0.01 | 0.77 | 0.77 | 0.01 | 0.01 |
| Kenya | 0.97 | 0.22 | 0.75 | 0.06 | 0.16 | 0.75 |
| Lesotho | 0.00 | 0.00 | 0.00 | 0.00 | 0.00 | 0.00 |
| Liberia | 0.56 | 0.02 | 0.54 | 0.54 | 0.01 | 0.01 |
| Madagascar | 0.52 | 0.24 | 0.28 | 0.04 | 0.28 | 0.20 |
| Malawi | 0.82 | 0.47 | 0.35 | 0.04 | 0.35 | 0.43 |
| Mali | 0.56 | 0.02 | 0.54 | 0.54 | 0.01 | 0.01 |
| Mauritania | 0.16 | 0.01 | 0.15 | 0.15 | 0.00 | 0.00 |
| Mozambique | 2.00 | 0.23 | 1.77 | 0.06 | 1.77 | 0.17 |
| Namibia | 0.10 | 0.02 | 0.07 | 0.01 | 0.07 | 0.02 |
| Niger | 0.29 | 0.04 | 0.25 | 0.25 | 0.01 | 0.03 |
| Nigeria | 4.32 | 0.22 | 4.10 | 4.10 | 0.10 | 0.13 |
| Rep. of the Congo | 0.25 | 0.07 | 0.18 | 0.18 | 0.02 | 0.05 |
| Rwanda | 0.27 | 0.05 | 0.23 | 0.02 | 0.02 | 0.23 |
| Sao Tome | 0.09 | 0.01 | 0.08 | 0.08 | 0.00 | 0.01 |
| Senegal | 0.85 | 0.03 | 0.82 | 0.82 | 0.01 | 0.02 |
| Sierra Leone | 0.67 | 0.03 | 0.65 | 0.65 | 0.01 | 0.01 |
| Somalia | 0.84 | 0.12 | 0.72 | 0.06 | 0.06 | 0.72 |
| South Africa | 0.50 | 0.12 | 0.38 | 0.05 | 0.38 | 0.07 |
| Sudan | 0.69 | 0.30 | 0.39 | 0.22 | 0.08 | 0.39 |
| Tanzania | 4.56 | 0.80 | 3.76 | 0.11 | 0.68 | 3.76 |
| Togo | 0.28 | 0.01 | 0.26 | 0.26 | 0.00 | 0.01 |
| Uganda | 0.80 | 0.17 | 0.62 | 0.08 | 0.09 | 0.62 |
| Zambia | 0.49 | 0.33 | 0.16 | 0.05 | 0.27 | 0.16 |
| Zimbabwe | 0.45 | 0.17 | 0.28 | 0.05 | 0.28 | 0.12 |

**Table S2**. Expected number of outbreaks caused in additional countries within one year by country of introduction.

| | **Predicted Positive** (Present) | **Predicted Negative** (Absent) |
|---|---|---|
| **Actual Positive** (Present, assuming clonal) | 237 | 81 |
| **Actual Negative** (Not present, assuming clonal) | 132 | - |
| | **Recall** = 71% (SD = 6) | |
| | **Precision** = 60% (SD = 2) | |
| | **Predicted Positive** (Present) | **Predicted Negative** (Absent) |
| **Actual Positive** (Present, assuming clonal) | 237 | 81 |
| **Actual Negative** (Not present, assuming nonclonal) | 32 | - |
| | **Recall** = 71% (SD = 6) | |
| | **Precision** = 82% (SD = 2) | |

**Table S3**. Validation across downsampled partitions, each with 20% downsampling. Table shows recovery (inferred presence or absence) of detected lineages, recall, and precision assuming clonal outbreaks (top) or nonclonal outbreaks (bottom).

| Parameter | Interpretation |
|---|---|
| $z_{v,i,t}$ | Hidden state of lineage presence or absence |
| $y_{v,i,t}$ | Observed lineages from genomic sequencing |
| $\pi_{v,i,1}$ | Initial probability of presence |
| $\mathbf{\Phi}$ | Transition process |
| $\mathbf{\Psi}$ | Observation process |
| $\psi_{v,i,t}$ | Probability of observed data given hidden state |
| $\rho_{v,i,t}$ | Probability of lineage presence |
| $\lambda_{v,i,t}$ | Sequences attributable to lineage $v$ |
| $\lambda^{*}_{v,i,t}$ | Prevalence of lineage $v$ |
| $\alpha_{v,i,t}$ | Forward probability of presence |
| $\mathbf{A}$ | Transition matrix |
| $\Phi^{k,l}_{v,i,t}$ | Probability of transition from state $k$ to state $l$ |
| $\phi_{v,j,t}$ | Transmission rate from country $j$ to country $i$ |
| $c_{i,t}$ | Cases in country $i$ in year $t$ |
| $\xi_{i,j}$ | Connectivity between country $i$ and country $j$ |
| $\eta$ | Scaling factor for cases |
| $\kappa$ | Gravity constant |
| $\tau_d, \tau_r$ | Scaling factors for donor and recipient countries, respectively |
| $d_{j,i}$ | Euclidean distance (km) between centroids of country $j$ and country $i$ |
| $\zeta$ | Scaling factor for distance |
| $\omega_{j,i}$ | Spatial random effect |
| $\delta$ | Within-country persistence |
| $\gamma_{v,i,t}$ | Probability of introduction from outside of the continent |
| $\beta_{v,i,t-1}$ | Backward probability of presence |
| $c^{*}_{v,i,t}$ | Inferred cases attributable to each lineage |

**Table S4.** Parameters used in the HMM model and their interpretations.

| *Lineage* | ***Timing*** *Median (95% HPD)* | ***Location*** |
|---|---|---|
| AFR9 | 1989.9557 (1987.5766, 1992.0304) | Liberia (0.3%), Guinea (95.1%), Senegal (0.1%), Cote d'Ivoire (0.4%), Other (4.1%) |
| AFR10 | 1995.4727 (1993.5098, 1996.6191) | Tanzania (34.5%), Comoros (0.1%), Uganda (0.4%), Kenya (64.8%), Other (0.2%) |
| AFR11 | 2000.6530 (1999.4473, 2001.5061) | Mozambique (91.0%), Zimbabwe (7.4%), South Africa (0.5%), Other (1.1%) |
| AFR12 | 2008.6833 (2008.1568, 2009.2207) | Cameroon (60.5%), Niger (1.3%), Nigeria (38.3%) |
| AFR13 | 2006.6934 (2006.3616, 2006.9792) | Kenya (99.4%), Other (0.6%) |
| AFR15 | 2019.8098 (2018.9354, 2020.7803) | Malawi (15.2%), South Africa (83.1%), Zimbabwe (1.6%), Other (0.1%) |

**Table S5.** TMRCA estimates of introduction timing (median and 95% HPD) and location of *V. cholerae* lineages AFR9-AFR15.
